# Swiss public health measures associated with reduced SARS-CoV-2 transmission using genome data

**DOI:** 10.1101/2021.11.11.21266107

**Authors:** Sarah A. Nadeau, Timothy G. Vaughan, Christiane Beckmann, Ivan Topolsky, Chaoran Chen, Emma Hodcroft, Tobias Schär, Ina Nissen, Natascha Santacroce, Elodie Burcklen, Pedro Ferreira, Kim Philipp Jablonski, Susana Posada-Céspedes, Vincenzo Capece, Sophie Seidel, Noemi Santamaria de Souza, Julia M. Martinez-Gomez, Phil Cheng, Philipp P. Bosshard, Mitchell P. Levesque, Verena Kufner, Stefan Schmutz, Maryam Zaheri, Michael Huber, Alexandra Trkola, Samuel Cordey, Florian Laubscher, Ana Rita Gonçalves, Sébastien Aeby, Trestan Pillonel, Damien Jacot, Claire Bertelli, Gilbert Greub, Karoline Leuzinger, Madlen Stange, Alfredo Mari, Tim Roloff, Helena Seth-Smith, Hans H. Hirsch, Adrian Egli, Maurice Redondo, Olivier Kobel, Christoph Noppen, Louis du Plessis, Niko Beerenwinkel, Richard A. Neher, Christian Beisel, Tanja Stadler

## Abstract

Genome sequences from evolving infectious pathogens allow quantification of case introductions and local transmission dynamics. We sequenced 11,357 SARS-CoV-2 genomes from Switzerland in 2020 - the 6th largest effort globally. Using a representative subset of these data, we estimated viral introductions to Switzerland and their persistence over the course of 2020. We contrast these estimates with simple null models representing the absence of certain public health measures. We show that Switzerland’s border closures de-coupled case introductions from incidence in neighboring countries. Under a simple model, we estimate an 86 - 98% reduction in introductions during Switzerland’s strictest border closures. Furthermore, the Swiss 2020 partial lockdown roughly halved the time for sampled introductions to die out. Finally, we quantified local transmission dynamics once introductions into Switzerland occurred, using a novel phylodynamic model. We find that transmission slowed 35 – 63% upon outbreak detection in summer 2020, but not in fall. This finding may indicate successful contact tracing over summer before overburdening in fall. The study highlights the added value of genome sequencing data for understanding transmission dynamics.

**One Sentence Summary:** Phylogenetic and phylodynamic methods quantify the drop in case introductions and local transmission with implementation of public health measures.

## INTRODUCTION

SARS-CoV-2 genomes were collected at an unprecedented scale in 2020 (*1*) and have been extensively used to characterize transmission dynamics, in particular because genetic data contains information on the epidemiological relationships between cases. These genomic data enable the reconstruction of introductions and downstream transmission chains in the absence of contact tracing data (*2*). Where contact tracing data is available, this approach has been verified and has additionally helped with linking unassigned individuals to known transmission chains (*3, 4*).

Several methods have been successfully used to reconstruct transmission dynamics at the onset of the COVID-19 pandemic using genetic data. Phylogenetic approaches reconstruct pathogen phylogenies and calculate relevant statistics from them without fitting any further explicit models. For example, phylogenetic reconstructions were used to show that reduced lineage size and diversity coincided with national lockdowns during the early Irish and English epidemics (*5, 6*). In Switzerland, (*7*) linked regional super-spreading events to a dominant lineage in the city of Basel using a phylogenetic reconstruction. Phylodynamic studies, on the other hand, assume the phylogeny arises from an underlying model of transmission between hosts, possibly including additional complexities like migration of hosts between regions. This assumption enables estimation of population-level transmission dynamics from pathogen genome data. For example, (*8–10*) showed that public health measures reduced SARS-CoV-2 transmission rates in Israel, New Zealand, and Washington State, USA.

New models and careful considerations of potential biases are required to quantify the effects of different public health measures in different regions. Here, we developed an analysis framework to quantify the association between the implementation and lifting of major public health interventions, such as border closures, lockdown measures, and contact tracing – three front-line tools in the fight against COVID-19 in 2020 – on transmission dynamics. Our framework uses a two-step process that carefully combines phylogenetic and phylodynamic methods to address potential sampling biases and phylogenetic uncertainty. Within the Swiss SARS-CoV-2 Sequencing Consortium (S3C; (*11*)) we sequenced 11,357 Swiss SARS-CoV-2 genomes until 1 December 2020. After combining these genomes with additional data available on GISAID (*12*) and down-sampling to control for biases in sampling efforts over time and among geographic regions, we were left with 5,520 Swiss SARS-CoV-2 genomes, representing up to 5% of weekly confirmed cases in Switzerland. We use these genomes to characterize transmission dynamics in Switzerland until the emergence and widespread dissemination of more transmissible variants of concern, starting in December 2020 (*13*). Our framework allows us to identify a clear effect of border closures and the spring 2020 partial lockdown on the rate of new introductions to Switzerland and their persistence. Furthermore, we were able to quantify the degree to which local transmission slowed upon outbreak detection. We find that this effect was strongest during summer 2020, when cases were low and contact tracing efforts likely more effective. To demonstrate the broader applicability of our analysis framework, we additionally analyzed data from New Zealand, where quarantine measures were stricter and local transmission was extremely limited throughout 2020. In New Zealand, we quantify a stronger transmission slowdown after outbreak detection, consistent with contact tracing there being highly effective.

## RESULTS

### Introductions and their persistence shed light on the effects of border closure and lockdown

First, we identified putatively independent introductions of SARS-CoV-2 into Switzerland and estimated their persistence. To do this, we selected SARS-CoV-2 genome sequences corresponding to up to 5% of confirmed cases each week, stratified to be geographically representative when possible (Figure S1). We divided these sequences by Pango lineage, as these lineages should represent monophyletic clades in the global SARS-CoV-2 phylogeny (*14*). Because of the hierarchical nature of Pango lineages, we aggregated lineages dominated by Swiss sequences into their respective parent lineages, allowing us to assume each analyzed lineage originated outside Switzerland (Table S1). To provide global context, we additionally selected the most genetically similar sequences from abroad for each lineage. We then constructed an approximate maximum-likelihood phylogeny for each such lineage of Swiss and genetically similar foreign sequences. We subsequently identified putatively independent introductions into Switzerland from these phylogenies, while allowing for a fixed number of export events. Importantly, we identified two plausible sets of introductions into Switzerland resulting from two different assumptions about the ordering of transmission events at polytomies with both Swiss and non-Swiss descendants. The set of “few” introductions was generated assuming the majority of polytomic lineages are from within-Switzerland transmission, whereas the set of “many” introductions was generated assuming the majority are new introductions. Sensitivity analyses show these two polytomy assumptions capture most of the uncertainty in the size and number of introductions amongst analyzed sequences (Supplementary text S1; Figure S2). Using additional data on which cases were from managed isolation and quarantine facilities in New Zealand versus identified in the community, we show that, as expected, the “many introductions” polytomy assumption is more realistic when the probability of infection abroad is high compared to the probability of locally acquired infection (Supplementary text S2). Throughout, we report uncertainty based on the difference between the few and many introductions sets.

We estimate that the analyzed sequences originate from between 557 (few) and 2284 (many) introductions into Switzerland. These introductions are roughly power law-distributed in size (Figure S3), with the 10 largest introductions accounting for 16 to 30% of sampled genomes. Introductions that yielded more than one sampled Swiss case in our dataset tended to be geographically constrained. Between 64% (few) and 92% (many) of sampled transmission chains (introductions with >1 sample) were sampled in only 1-2 of the 26 Swiss cantons (Figure S4A). As expected, larger introductions were sampled in more cantons (Figure S4B; Pearson’s R between introduction size and number of cantons is 0.86 for many introductions, 0.75 for few introductions). From a down-sampling analysis, we observe that if we were to include more sequences, we would identify more introductions (Figure S2C). Therefore, the analyzed genomes do not represent all introductions into Switzerland but, given the samples are spatio-temporally representative, are a representative subset of introductions. Due to incomplete sampling, each sampled introduction contains only a subset of all cases in the full transmission chain.

Since we sampled sequences proportionally to confirmed cases through time (Figure S1A; R^2^ between number of confirmed cases and number of analyzed Swiss sequences each week 0.72), we can assume that trends through time in the number and persistence of introductions are representative of the underlying dynamics. Figure 1A shows the number of newly sampled introductions identified each week from our dataset, which peaked the week of 15 March under both polytomy assumptions. Switzerland closed its external borders to Italy 13 March 2020 and with the rest of the world shortly thereafter (*15*). To disentangle the effect of the border closures versus local control measures, we back-calculated the expected number of total (both sampled and unsampled) introductions each week under a birth-death skyline model (*16*). This calculation corrects for the probability that an introduction went extinct or remained unsampled each week until the end of the sampling period, given estimates of the sampling proportion and the time-varying effective reproductive number Re in Switzerland. Then, we develop a simple null model that assumes that prior to 13 March 2020, total introductions are a linear function of case counts in Switzerland’s largest neighboring countries (Italy, France, Germany, and Austria). Here we are assuming incidence in travelers to Switzerland follows incidence in the general community in these countries. Figure 1C shows this model fit to total introduction estimates generated based on each polytomy assumption and model projections (dashed lines) from 13 March through the partial re-opening of Switzerland’s European borders on 15 June 2020 (*15*). In the following, we report uncertainty based on the 95% HPD upper and lower bound estimates for Re used to estimate total introductions. Uncertainty in travel patterns is discussed later. Compared to the null model, we estimate a reduction of 7,000 (few introductions; uncertainty 4,500 - 11,000) or 79,000 case introductions (many introductions; uncertainty 41,000 - 130,000). Despite the high uncertainty in the absolute number of introductions averted depending on the polytomy assumption and the precise value of Re in Switzerland, we estimate a consistent percentage-wise reduction of 94.1% (few introductions; uncertainty 85.9 - 97.8%) or 94.2% (many introductions; uncertainty 86.2 - 97.9%). We note that total European case counts peaked later than in Switzerland’s neighboring countries while our analysis only considers neighboring countries. Thus, the period of high import pressure may have extended longer than we assume, depending on where most introductions were coming from (Figure S5). However, our focus on neighboring countries is supported by travel statistics. For instance, neighboring countries comprise 99% of cross-border working permits granted by Switzerland for the first quarter of 2020 (approximately 330,000 individuals). These countries also account for 36% of registered arrivals at Swiss hotels in January and February 2020 (approximately 450,000 individuals) (*17*). Thus, we assume introduction dynamics are largely driven by these neighboring countries. However, our estimates of the precise reduction in imported cases depend strongly on this assumption.

**Fig. 1.**
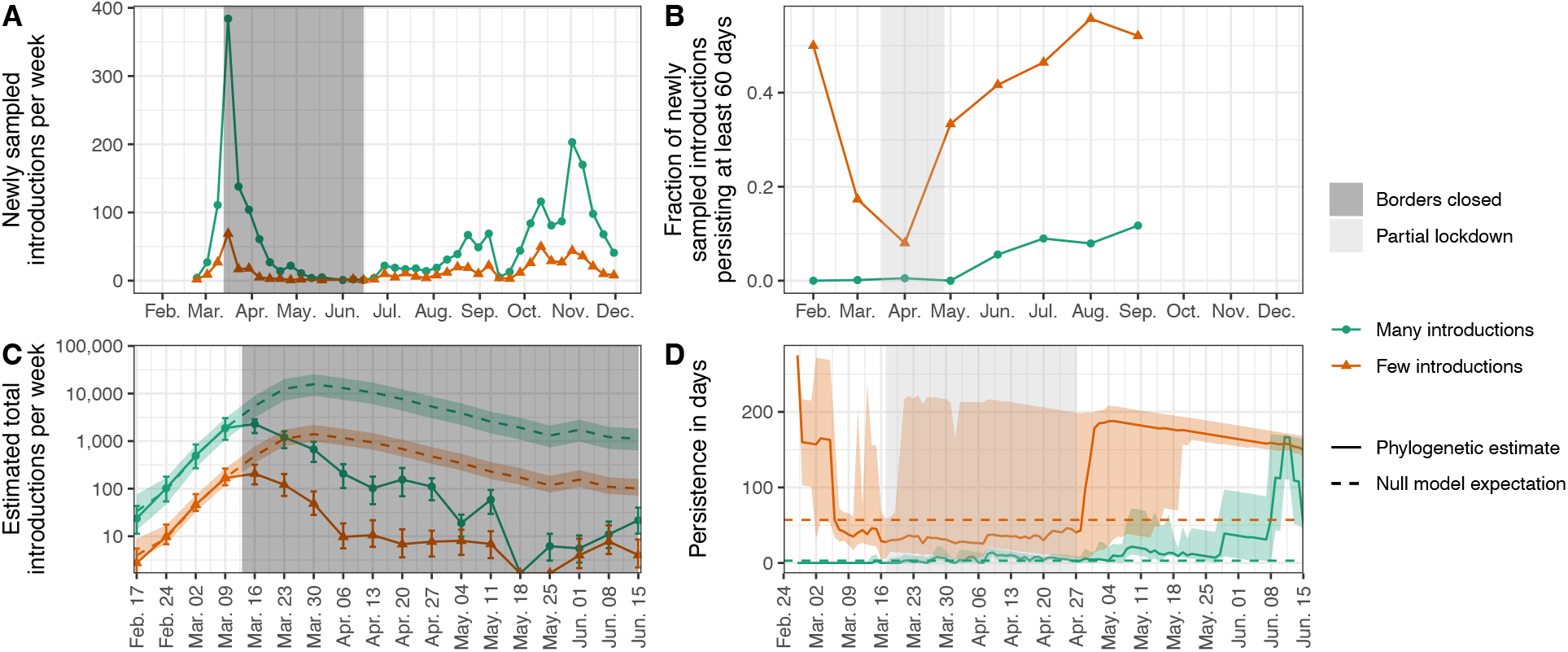
Genome-based estimates of SARS-CoV-2 introductions into Switzerland and their persistence. (**A**) shows the number of newly sampled introductions identified each week and (**B**) shows the fraction of newly sampled introductions each month that persist for at least 60 days from the oldest to the most-recent sample. This persistence measure is only defined until September because we only considers sequences obtained until 1 December 2020. Orange and green correspond to estimates generated under the few and many introductions polytomy assumptions, respectively. (**C**) and (**D**) focus on dynamics around the Swiss border closure and partial lockdown periods, which are highlighted with shaded rectangles. (C) shows estimated total introductions (solid lines) compared to a null model (dashed lines) where total introductions are a linear function of case numbers in Switzerland’s neighboring countries. The null model is fit to the points prior to the border closure, values after that are projections. Uncertainty bounds for total introductions (error bars) and null model predictions (colored shaded areas) are based on the 95% upper and lower HPD bounds for Re when estimating total introductions. Uncertainty in travel patterns is not shown, see Figure S5. (D) shows the distribution of ongoing persistence for introductions circulating each day (solid lines), compared to a null model (dashed lines) where persistence is constant through time (equal to the median calculated until 15 June). Solid lines are median time to last sampling amongst introductions newly sampled or still ongoing each day. The shaded areas show the interquartile range of this persistence distribution.

New introductions cannot sustain an epidemic unless they persist in the local population. Our analysis suggests several introductions were quite persistent in Switzerland, including one that may have persisted across our entire sampling period (Figure S6). On average, introductions persisted 5 days (many introductions; standard deviation 16 days) to 34 days (few introductions; standard deviation 53 days) from the oldest to the most-recent sample of each introduced lineage in our dataset. Lineage persistence until last sampling was lower during the partial lockdown (17 March - 27 April; Figures 1B and D) compared to summer 2020. While only 0.5 - 8% of introductions in April were sampled for at least 60 days, this fraction increased to 12 - 52% in September, just before a large fall wave in Switzerland. We also developed a simple null model to assess whether the spring 2020 lockdown measures and associated behavioral changes affected the persistence of introduced lineages. Here, our null model is that persistence, measured as the time until introductions circulating each day are last sampled, does not change through time. We assume this delay distribution always equals the median persistence calculated over the spring period (until 15 June). Figure 1D contrasts this null model assumption with empirical persistence calculated from each day under each polytomy assumption. The distribution does indeed vary through time, deviating from the null model. We estimate median persistence of introductions at the start of the lockdown is less than or around the median calculated over the whole spring and rises to above this null model threshold in the post-lockdown period. Quantitatively, introductions persisted roughly twice as long until last being sampled at a post-lockdown peak around 10 June compared to at the lockdown start (Figure 1D). We note that under the few introductions assumption, persistence estimates are upper-bounded by the end of our sampling period, so the increase in persistence may also be an underestimate (Figure 1D).

### Phylodynamic model indicates summer introductions slowed after detection

Next, we investigated local transmission dynamics once SARS-CoV-2 lineages were introduced to Switzerland in more detail. To do this, we quantified time-varying transmission dynamics in Switzerland in a Bayesian phylodynamic framework. As a base model, we used the birth-death model with serial sampling originally described in (*18*). We modified the model to condition on the previously identified few or many introductions sets, i.e., sequences from each introduction have an independent origin. In a nutshell, the model assumes that once lineages are introduced, they are (i) transmitted between hosts, according to a time-varying transmission rate which is the same across all introductions; (ii) die out upon recovery/death of the host, according to a constant becoming-uninfectious rate; and (iii) yield genome samples with a time-varying sampling proportion which is the same across all introductions. We assume individuals who test positive adhere to self-isolation regulations, so sampling corresponds to a death event for the viral lineage. Under this parameterization, Re is a function of the transmission rate, becoming-uninfectious rate, and sampling proportion.

We developed a novel extension to this methodology by adding a transmission rate “damping” factor, as shown in Figure 2. The transmission rate is allowed to decrease by a multiplicative damping factor two days after an introduction is first sampled. We use a spike-and-slab prior on this factor to include the possibility of no transmission slowdown. We allow this damping factor to vary between spring, summer, and fall 2020 - periods characterized by very different case numbers and testing regimes in Switzerland (Figure 3A; (*19*)). Using this model, we aim to test whether contact tracing efforts in Switzerland slowed transmission once introductions were detected. We reason that test-trace-isolate can only slow transmission from shortly after the first case of an introduction tests positive but not beforehand, as beforehand the introduction was circulating cryptically. The two-day delay aims to account for the time between an individual giving a sample (i.e., being swabbed) and having their contacts notified.

**Fig. 2.**
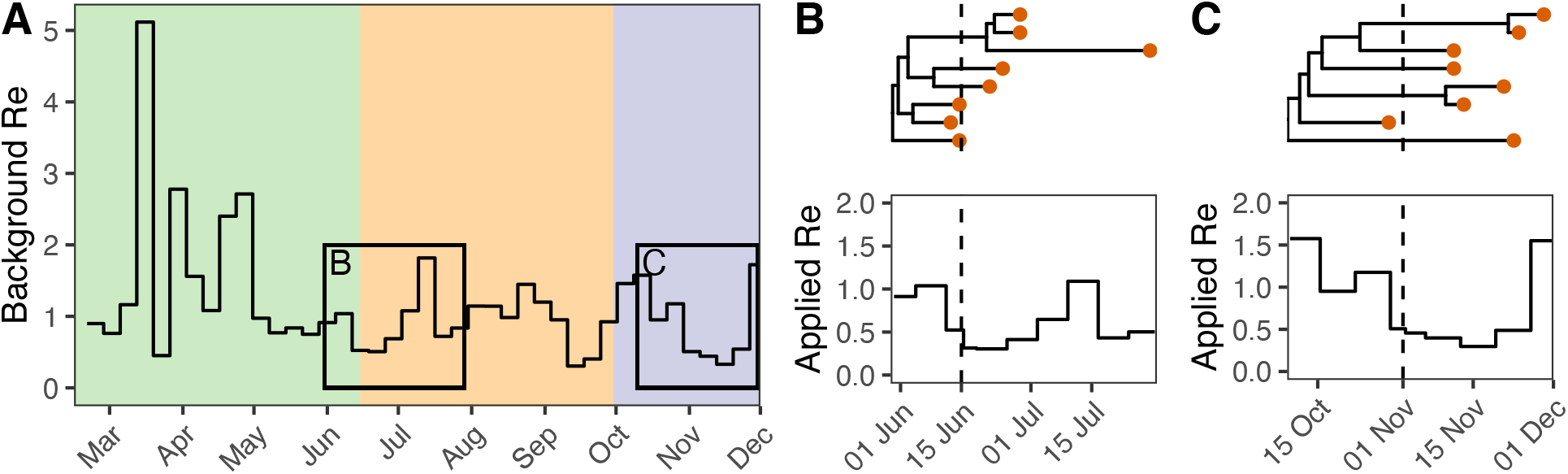
Illustration of how transmission rate damping is modeled. (**A**) shows a background Swiss-wide time-varying effective reproductive number Re before any damping. Here we show the median posterior result from the model applied to the many introductions data as an illustration. In each of the colored areas (green = spring, orange = summer, and purple = fall), a different damping factor is proposed. The black boxes in (A) highlight the spread of two real introductions (**B**) and (**C**) generated under the many introductions polytomy assumption. The genome data sampled from these introductions are shown as red dots in (B) and (C). The appropriate damping factor on Re is applied to each introduction 2 days after the first genome sample (dashed lines). We used 0.6 for the summer damping factor and 0.9 for fall for this illustration. The likelihood of the genome sequence data at the tips of the phylogenies is calculated given the “applied” Re specific to each introduction (B and C, bottom).

**Fig. 3.**
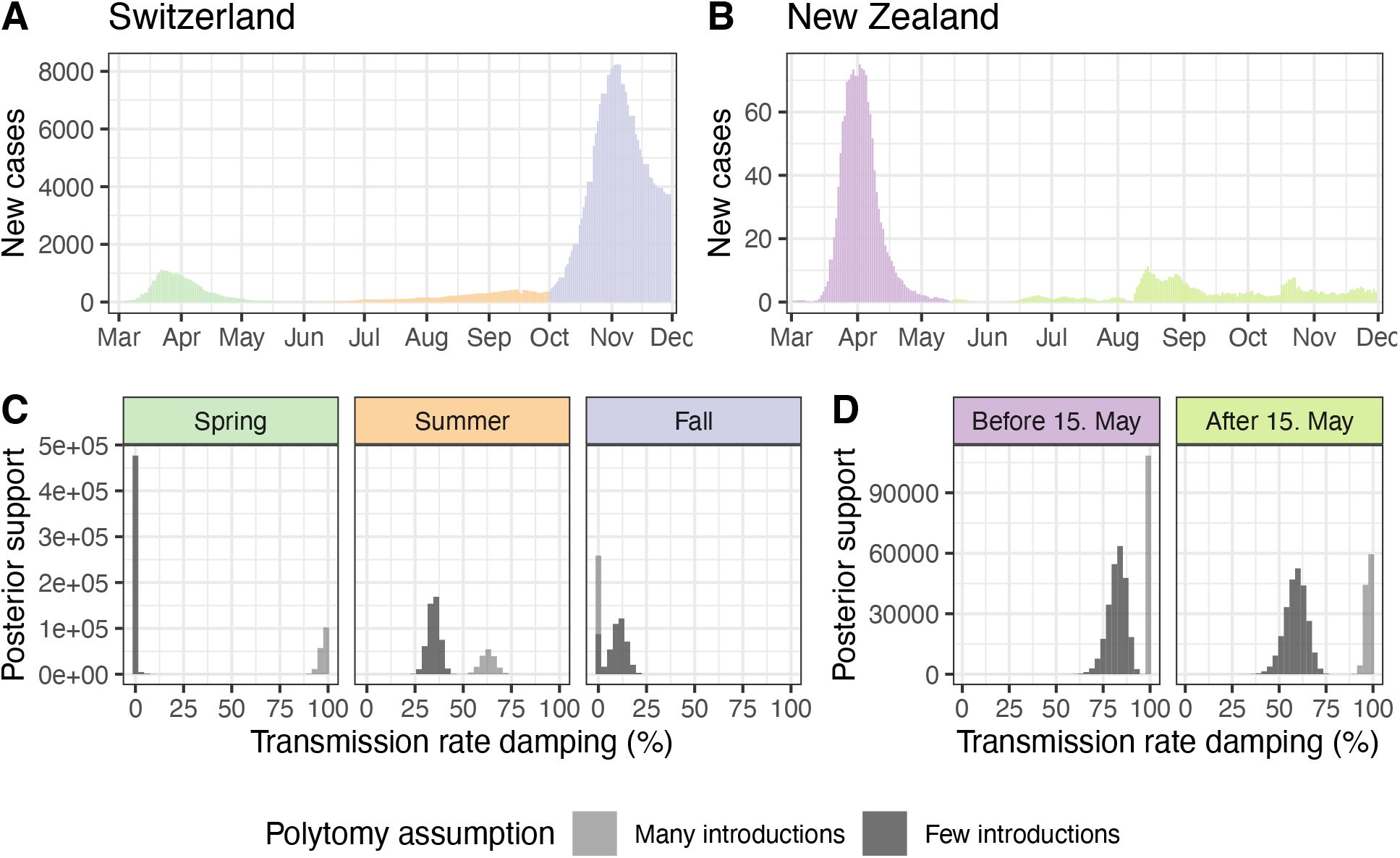
Phylodynamic estimates for the transmission damping factor in Switzerland and New Zealand compared to case numbers. Case numbers in (**A**) Switzerland and (**B**) New Zealand during 2020 are shown as a 7-day rolling average of daily new confirmed cases (22). (**C**) and (**D**) show estimates for if and how much transmission rates were dampened after introductions were sampled during different time periods in (C) Switzerland and (D) New Zealand. The inference was done twice, once conditioning on introductions identified assuming many introductions (light gray) and once assuming few introductions (dark gray). Thus, the difference between estimates in light and dark gray are due to phylogenetic uncertainty. Results shown are from the model with an unbounded sampling proportion prior, results with a bounded sampling proportion prior are similar (Figure S8).

Specifically, this delay consists of the time to RT-PCR results, which was generally below 24 hours in Swiss diagnostic laboratories (*20*), plus the time for contact tracers to reach contacts or an individual to receive and input their positive test code to the SwissCovid contact tracing app. We fit the phylodynamic model in several configurations: conditioning on either the many or few introductions set, using a bounded or an unbounded sampling proportion prior (see Supplementary text S3), and with or without a transmission damping factor.

Across these model configurations, we recover roughly the same trends in Re as estimates based on confirmed case numbers beginning with the first analyzed sequence from 27 February (Figure S7B). Compared to confirmed case-based estimates, we estimate a sharper decline in Re coinciding with lockdown measures. Depending on the polytomy assumption, we estimate Re was 2.2 (many introductions; 95% HPD 1.5 - 2.9) or 3.5 (few introductions; 95% HPD 2.9 - 4.2) in the week of 9 March. Re fell to 0.3 (many introductions; 95% HPD 0.2 - 0.4) or 0.4 (few introductions; 95% HPD 0.2 - 0.6) in the week of 16 March 2020 (posterior median estimates with no damping factor and an unbounded sampling proportion prior). With a bounded sampling proportion, peak Re estimates are slightly higher (Figure S7). Results in fall 2020 are highly dependent on the sampling proportion prior, where Re estimates better match confirmed case-based estimates when the sampling proportion is treated as a fitting parameter (i.e., with an unbounded prior, resulting in unrealistic estimates of the sampling proportion; see Figure S7A).

From the model fit with a damping factor, we estimate a 35% (few introductions; 95% HPD 29 - 41) - 63% (many introductions; 95% HPD 56 - 70) slowdown in transmission after introductions are first sampled in summer 2020 (posterior median estimates with an unbounded sampling proportion prior). In comparison, there is little support for a slowdown effect upon the first sampling during fall 2020 (Figure 3). These results are qualitatively robust to bounding the sampling proportion prior (Figure S8). In contrast, damping factor estimates in spring 2020 are inconsistent, depending on the polytomy assumption. Low genomic diversity in SARS-CoV-2 during this period causes high phylogenetic uncertainty (*21*; see also the differences in several selected introductions in Figure S9). This results in quite different estimates for the damping factor depending on the polytomy assumption used. In summary, we report a summer 2020 “slowdown” dynamic in SARS-CoV-2 transmission in Switzerland, where transmission slows after the first genome in a new introduction is sampled. This slowdown is not observed in fall 2020.

### New Zealand data shows slowdown effect is not Switzerland-specific

While Switzerland is centrally located in Europe and well-connected to other countries, especially those in the (normally) barrier-free Schengen zone, New Zealand is a relatively isolated island nation. Additionally, New Zealand aimed to eradicate SARS-CoV-2 throughout 2020 using strong measures, such as keeping its borders closed and enforcing strict quarantine-on-arrival (*23*), while Switzerland partially reopened its borders to Europe on 15 June. We applied the same analysis framework for introduction estimation and phylodynamic inference to SARS-CoV-2 sequences from New Zealand as a comparison to our Switzerland-specific results. For the phylodynamic analysis, we estimated independent damping factors before and after an epidemic breakpoint in mid-May 2020 when local transmission was briefly eradicated (*9, 24*). Case numbers were subsequently held at lower levels through December 2020 (Figure 3B, *24*).

From the model fit with a damping factor, we estimate transmission damping in New Zealand before and after 15 May to be comparable with or stronger than in Switzerland during summer and fall 2020 (Figure 3D), regardless of the polytomy assumption used. Thus, the existence of a transmission damping effect is not specific to Switzerland. From the model fit without a damping factor, our estimates for the sampling proportion and Re are inconsistent. In particular, the sampling proportion is estimated to be unrealistically high when conditioning on the many introductions data set. However, including the damping factor in the model reconciles estimates based on each polytomy assumption, yielding more realistic estimates for the sampling proportion and pre-damping Re (Figure S10).

## DISCUSSION

We quantify the change in cross-border and local transmission dynamics with the introduction or lifting of major public health measures in Switzerland based on genome sequence data. First, we quantify the reduction in case introductions during the period of Switzerland’s strictest border closures. Travel from Italy was tightly restricted beginning 13 March and with the rest of the world beginning 16 March 2020. These measures were partially lifted on 15 June, when Switzerland re-opened to European countries in the Schengen zone (*15*). We used phylogenetic estimates for the number and timing of viral introductions into Switzerland to show that newly sampled introductions peaked during the week of 15 March, coinciding with the implementation of border closures. Due to many identical or near-identical SARS-CoV-2 lineages circulating widely in Europe during spring 2020, the total number of introductions to Switzerland is highly uncertain. We considered two extreme cases, encompassing most of the phylogenetic uncertainty in the size and number of introductions. We additionally corrected these estimates based on the time-varying probability that an introduction went unsampled. After disentangling the effect of border closures and local control measures in this way, we show that border closures de-coupled introduction dynamics from case counts in neighboring countries. Compared to a simple null model assuming that the incidence in travelers corresponds to the incidence in Switzerland’s neighboring countries, we quantify an 86 - 98% reduction in case imports from 13 March – 15 June. While the de-coupling of case introductions and incidence in neighboring countries is clear, our estimates for precisely how many and what fraction of introductions were averted are subject to several strong assumptions, namely that incidence in travelers is the same as the average in the different source populations, and that the majority of imported cases would have come from Switzerland’s neighboring countries. Finally, we note that the fraction of polytomic lineages that were independent introductions likely decreased throughout spring 2020 as local incidence rose, travel declined, and the probability of locally acquired infection rose (Supplemental text S2). We expect the truth to lie somewhere between the estimates generated under our two polytomy assumptions.

Second, we quantify the reduction in local transmission during Switzerland’s partial lockdown in spring 2020 compared to the pre- and post-lockdown time period. A suite of lockdown measures, including closure of schools, non-essential shops, restaurants, and entertainment and leisure establishments was introduced on 17 March 2020. Many non-essential shops re-opened on 27 April, before schools and most other shops reopened on 11 May (*25*). We estimate that sampled introductions circulating on 17 March persisted only about half as long until last sampling as in mid-June. We also estimate that only 0.5 - 8% of newly sampled introductions in April persisted more than 60 days until last being sampled, compared to 12 - 52% in September. These findings agree with previous findings (*26*), which demonstrated a reduction in the number of transmission clusters and the risk of transmission within clusters in the Canton of Vaud, Switzerland after the implementation of lockdown measures. Finally, we obtained genome-based estimates for the time-varying effective reproductive number Re in 2020 from our phylodynamic model. We estimate that Re dropped from 2.2 - 3.5 the week of 9 March to 0.3 - 0.4 the week of 16 March, coinciding with lockdown measures. Two models fit to hospitalization and death (*27*) and confirmed case (*28*) data in Switzerland gave similar or slightly lower pre-lockdown Re estimates of 2.1 - 3.8 and 1.6 - 1.9, respectively, and similar or slightly higher post-lockdown Re estimates of 0.3 - 0.6 and 0.6 - 0.8 after 29 March, respectively. Our phylodynamic estimates, which account for an influx of introduced cases, suggest a sharper reduction in Re coinciding with the Swiss lockdown than these estimates based on epidemiological data. This could be due to accounting for imported cases or the case-count smoothing used by (*27, 28*).

Finally, we quantified a summertime “slowdown” dynamic in Switzerland in which introductions initially spread faster, then slowed 35 - 63%. This dynamic was not observable in fall 2020 in Switzerland. A plausible explanation of this dynamic is a successful test-trace-isolate implementation that roughly halved transmissions once an introduction was identified during summer 2020 in Switzerland. We cannot make a statement about the relative speed of transmission chains pre- and post-first sampling in spring 2020. This is because many lineages are ambiguous as to whether they were imported and died out quickly, or resulted in extensive, ongoing local transmission. Therefore, conditioning the birth-death phylodynamic model on few or many introductions during this period yields very different results. For the damping factor analysis, we make the strong assumption that transmission in all lineages descending from an introduction slows simultaneously 2 days after the first genome sample belonging to the introduction is collected. This may be justified if efficient informal backward contact tracing occurred or if individuals in sister lineages were identified around the same time but their samples were not sequenced or not included in our analysis. Then, there are other possible explanatory factors at play. First, travelers returning to Switzerland during summer 2020 have been implicated in transmitting more than non-travelers (*29*). Thus, a passive transmission slowdown might have happened as introduced lineages moved into the non-traveler population. We would expect travelers in fall to have similar contact networks as those in summer, but we do not quantify a transmission slowdown in Switzerland in fall. This coincides with high case numbers during a fall wave, when Swiss contact tracing was reported to be overburdened (*30*). Second, contacts of positive cases are likely tested more intensely, potentially yielding “bursts” of samples around the first detected cases that subsequently disappear. If so, we can still interpret the slowdown dynamic as evidence that test-trace-isolate implementation was working, but it is difficult to determine precisely by how much transmission actually slowed.

International comparisons also lend perspective to the transmission slowdown effect we quantify from Swiss genome data. Using the same analysis framework, we quantified a significant slowdown effect in New Zealand during two different time periods. Thus, this slowdown effect is not unique to Switzerland in summer 2020. Importantly, (*4*) showed - using genome sequence data - that New Zealand contact tracing was highly effective in identifying SARS-CoV-2 infection clusters. Then, (*31*) exploited an accidental, partial breakdown of English contact tracing to show that normal contact tracing in early fall 2020 reduced transmissions by 63% in the 6 weeks following a positive case. This measure is within the range of our estimates for a transmission slowdown in Switzerland in summer 2020.

Together, our results quantify the reduction of case importation and local transmission in Switzerland during the spring 2020 partial lockdown and partial border closure periods. Further, we provide genome-based quantification of a summertime transmission slowdown in Switzerland that may be linked to successful contact tracing efforts. This slowdown is not observed in fall when contact tracing efforts were overwhelmed in Switzerland but is observed in data from New Zealand in 2020. We have shown that our inference framework is straightforward to apply to different datasets and produces quantitative estimates that we envision can help policy-makers weigh general and specific measures against the respective burdens they impose.

## MATERIALS AND METHODS

### Genomic surveillance by the Swiss SARS-CoV-2 Sequencing Consortium in 2020

Altogether 11,357 SARS-CoV-2 genome sequences sampled in Switzerland during 2020 were generated by the Swiss SARS-CoV-2 Sequencing Consortium (*11*). This sequencing effort represents the majority (79%) of Swiss SARS-CoV-2 genome sequences collected in 2020 and represents the 6^th^ largest contribution of SARS-CoV-2 sequences globally in 2020 (Table S2), based on data available on GISAID as of June 2022 (https://www.gisaid.org/; (*12*)). Here, we briefly describe how these samples were generated.

RNA extracts from qPCR-positive patient nasal or oropharyngeal swabs were provided by Viollier AG, a Swiss medical diagnostics company. RNA was extracted using either the Abbott m2000sp or Seegene STARMag 96×4 Universal Cartridge kits. Extracts were then transferred to the Genomics Facility Basel or the Functional Genomics Center Zurich for whole-genome sequencing. Both centers used the ARTIC v3 primer scheme (*32*) to generate tiled, approximately 400bp-long amplicons. Library preparation was done with the New England Biolabs (NEB) library preparation kit. Libraries were sequenced on Illumina MiSeq or NovaSeq machines, resulting in 2 × 251 basepair reads. Bioinformatics processing was performed using V-pipe (*33*), including read trimming and filtering with PRINSEQ (*34*), alignment to GenBank accession MN908947 (*35*) with bwa (*36*), and consensus base calling. Positions with <5x coverage were masked, positions with >5% and >2 reads supporting a minor base were called with IUPAC ambiguity codes, and positions with >50% reads supporting a deletion were called as a deletion. We rejected samples with <20,000 non-N bases. The consensus sequences are available in the Global Initiative on Sharing Avian Influenza Data (GISAID) repository (*12*) under submitting lab “Department of Biosystems Science and Engineering, ETH Zürich”.

### Dataset construction and sampling procedure

From all sequences available on GISAID (accessed 31 May 2021), we filtered the collection date to on or before 1 December 2020, removed non-human sequences, and sequences <27,000 bases long. We also filtered sequences flagged by the Nextclade tool (*37*) for suspiciously clustered SNPs (QC SNP clusters status metric not “good”; >= 6 mutations in 100 bases), too many private mutations (QC private mutations status metric not “good”; >= 10 mutations from the nearest tree node), or overall bad quality (Nextclade QC overall status “bad”). We aligned sequences to the reference genome MN908947.3 using MAFFT (*38*). Finally, we followed the Nextstrain pipeline’s recommendation to mask the first 100 and last 50 sites of the alignment (*39*) since the start and end of SARS-CoV-2 sequences are prone to sequencing errors (*40*).

From all available Swiss sequences, we sampled up to 5% of confirmed case counts in each Swiss canton each week until 1 December 2020. Confirmed case data was provided by the Swiss Federal Office of Public Health (now available on https://www.covid19.admin.ch) (Figure S1). At the time of data access, cases were only attributed at the cantonal level beginning in mid-May. Before then, we sampled randomly from across Switzerland. Where not enough sequences were available from a canton in a week, we used all available sequences. To reduce the size of the alignments for phylogenetic analysis, we divided the focal Swiss set into Pango lineages (*14*), similar to (*10*). Lineages composed of >50% Swiss sequences were aggregated into their parent lineage(s) until <= 50% were Swiss. This aims to ensure that each analyzed lineage originated outside of Switzerland. Table S1 lists the analyzed aggregated lineages and the number of sequences per lineage.

We then added the most genetically similar sequences from abroad to each lineage alignment to add a global context. This aims to help distinguish between SARS-CoV-2 variants unique to Switzerland (likely within-Switzerland transmission) and variants also circulating abroad (possibly recent introductions or exports). We considered all non-Swiss sequences from each lineage available on GISAID that pass the quality filtering steps detailed above and applied the Nextstrain priority script (*39*) to rank these sequences by their genetic similarity to Swiss sequences in each lineage alignment. Briefly, the priority script ranks a set of foreign context sequences by the Hamming distance to their nearest neighbor within a set of focal sequences. Context sequences are further penalized for having high numbers of masked positions or for being more distant neighbors of the same focal sequence. We selected twice as many context sequences as focal Swiss sequences for each analyzed lineage alignment. Our results are based on a final set of 5,520 focal sequences from Switzerland and 11,009 genetically similar sequences from abroad, which were divided into 148 lineage alignments (Table S1).

### Phylogenetic analysis

We estimated an approximate maximum likelihood phylogeny for each lineage alignment using IQ-TREE (*41*) under an HKY substitution model (*42*) with empirical base frequencies and four gamma rate categories to account for site-to-site heterogeneity (*43*). We added one of the earliest collected SARS-CoV-2 genomes Wuhan/WH01/2019 (GISAID strain EPI_ISL_406798, GenBank accession MT019529.1) as an outgroup for rooting to each alignment and estimated branch lengths in calendar time units using least-squares dating (LSD) (*44*) implemented in IQ-TREE. We used a strict molecular clock and a minimum mutation rate of 8 × 10^−4^ substitutions per site per year (s/s/y), based on estimates by Nextstrain (*45*). We constrained the most-recent common ancestor to be between 15 November and 24 December 2019, also based on estimates by Nextstrain (*45*), and set the minimum branch length to zero. Sequences that violated the strict clock assumption (Z-score threshold > 3) were removed and near-zero length branches (< 1.7 × 10^−5^ substitutions per site) were collapsed into polytomies, reflecting the fact that the sequence data alone is not sufficient to resolve the ordering of these transmission events. Given the root date constraints, the mutation rate conformed to the lower bound of 8 × 10^−4^ with extremely narrow confidence intervals. After removal of sequences violating the strict clock assumption, 5,452 sequences remained across all lineage trees.

### Identifying introductions

We identified putative Swiss transmission chains (collections of two or more genome sequences resulting from within-Switzerland transmissions) from each lineage tree while allowing for a fixed number of export events. We used the following criteria applied on a recursive tip-to-root tree traversal: at least two Swiss sequences are part of a clade in the tree and the subtree spanned by these Swiss sequences is monophyletic upon removing (a) up to three export events where (b) only one export event may occur along each internal branch. Exports are clades containing non-Swiss sequences. We chose a conservative value for (b) while still allowing some exports and note that the number of inferred transmission chains is robust to different values for (a) given (b) (Figure S2A). We assume the identified transmission chains and remaining singleton Swiss sequences each represent an independent introduction into Switzerland.

We repeated this procedure twice for each lineage tree, making different assumptions upon reaching a polytomy where non-Swiss descendent(s) of the polytomy would cause the proposed introduction to violate criterion (a). First, we split all Swiss clades descending from the polytomy into independent introductions. The second time, we aggregated descendent Swiss clades, going in descending size order, into a single introduction. If in doing this we reached criterion (a), we continued aggregating descendants into a second introduction, and so on. The above procedures are heuristic, but analogous to the ACCTRAN (accelerated transformations) and DELTRAN (delayed transformations) methods for assigning character transformations when multiple scenarios are equally parsimonious (*46*). In summary, we identify introductions twice, generating estimates that represent two plausible sets of many and few introductions at polytomies, where sequence data is not informative about the order of the branching events.

### Uncertainty in identifying introductions

We evaluated the effect of several variables on the number and size of identified introductions, as discussed in Supplementary text S1. We found that our two different polytomy assumptions are sufficient to capture most of the uncertainty in the number and size of introductions due to the specific heuristic criteria used to identify introductions from a phylogenetic tree (Figure S2A). As expected, increasing the ratio of foreign context to focal Swiss sequences analyzed identifies more, smaller introductions compared to a lower ratio. However, our two different polytomy assumptions at a 2:1 ratio are again sufficient to capture most of this uncertainty (Figure S2B).

### Quantifying the reduction of introductions during the time of border closures

Prior to fitting our null model for introductions through time, we back-calculated the total number of introductions each week expected under a birth-death skyline model, as described in the section “Phylodynamic analysis” below. Under this model, one can calculate the probability *p(t)* that a new introduction at time *t* would have no sampled descendants by 1 December 2020. This formula is given in (*16*). We used weekly time bins, taking the median and 95% HPD upper and lower bounds for Re from our phylogenetic analysis (see below), a constant sampling proportion of 5% based on our known sampling scheme, and a constant become-uninfectious rate of 36.5 per year, which corresponds to an average of 10 days to becoming uninfectious (roughly in line with estimates provided by the Swiss Federal Office of Public Health (*47*)). We divided the number of sampled introductions each week by 1 - *p(t)*, the probability an introduction at the start of the week would yield a sampled descendant by 1 December 2020. This yields an estimate for the total number of introductions each week (both sampled and unsampled), while accounting for varying local transmission dynamics. For a more extended description of the implementation of this correction, see the Supplementary text S4.

Then, we assumed a simple null model in which introductions are a linear function of case counts in Switzerland’s largest neighboring countries: Italy, France, Germany, and Austria. We used a 7-day rolling average of case count data from the European Centre for Disease Prevention and Control (ECDC) (*22*). Further, we considered up to 18 days delay between the actual introduction event and an introduction being sampled. This is based on the 8-day lag from importation to first local transmission estimated by (*6*) in the U.K. and a 10-day infectious period. We back-calculated total introductions as described above for each plausible delay value using either the median or 95% upper or lower HPD Re estimate from our phylodynamic analysis (see below). We fit the model independently to each of these weekly estimates up to 13 March. We selected the delay yielding the best model fit (lowest root mean squared error using the median Re estimate) for each set of few or many introductions. These were 4 and 5 days, respectively. Finally, we projected introductions after 13 March using the fitted model coefficients and ongoing case counts in the surrounding countries. We did not fit the model to data after border closures were partially lifted because travel behavior was still affected by risk of infection, risk of new restrictions being introduced, and ongoing stay-at-home guidance. This is apparent in data collected by the Swiss Tourism Federation, which demonstrates a marked drop in overnight stays by foreign residents in Switzerland from approximately 6.3 million in the winter season November 2019 - April 2020 to 3.1 million in the summer season May - October 2020 (*48*). As a sensitivity analysis, we also fit the model using confirmed cases in all non-Swiss European countries as defined in the ECDC’s case count data (*22*) (Figure S5).

### Quantifying the reduction of persistence of introductions during the lockdown

We developed a second simple null model to test whether the Swiss partial lockdown from 17 March to 27 April 2020 coincided with a change in the persistence of introductions. This null model assumes that in the absence of measures, introductions circulating on any given day persist equally long. In other words, introductions die out (are no longer sampled) according to a delay distribution that is constant through time. For each date, we calculated the time from that date to the last sample for each introduction persisting on that date. Singleton introductions are trivially assumed to persist for 1 day. Then, we report the median and interquartile range of this delay distribution from each date.

### Phylodynamic analysis

After identifying introductions, we performed phylodynamic inference on them using the BDSKY (birth-death skyline) method (*16*) in BEAST2 (*49*). To avoid model mis-specification due to the more transmissible alpha variant, we analyzed data only until 1 December 2020. We also pruned introductions to only include genomes generated by the S3C, as these were explicitly surveillance samples. This left 4,136 genome sequences for phylodynamic analysis. The phylodynamic inference relies on two main models: a nucleotide substitution model describing an evolutionary process and a population dynamics model describing a transmission and sampling process. For the nucleotide substitution model, we assumed an HKY (*42*) model with four Gamma rate categories to account for site-to-site rate heterogeneity (*43*). We used the default priors for kappa and the scale factor of the Gamma distribution. We assumed a strict clock with the clock rate fixed to 8 × 10^−4^ s/s/y, as estimated by (*45*).

For the population dynamics model, we used BDSKY (*16*). In BDSKY, the identified introductions are the result of a birth-death with sampling process parameterized by an effective reproductive number, a becoming-uninfectious rate, and a sampling proportion. As in (*10*), we inferred these population dynamical parameters jointly from the different introductions. More concretely, each introduction is assumed to result from an independent birth-death process having its own origin time, but sharing all other parameters with the processes associated with the other introductions. We applied a uniform prior on the time of origin for each introduction, between 15 February and the oldest sample in the introduction. This constrains introductions to have an origin no earlier than 15 February, excluding the possibility of introductions and subsequent local transmission before the date the first confirmed Swiss case was reported infected abroad in Italy (*50*). After 15 February, our prior expectation is a uniform rate of introductions through time. We fixed the become-uninfectious rate to 36.5 per year, as above. We allowed Re to vary week-to-week, with an Ornstein-Uhlenbeck smoothing prior applied to the logarithm of this parameter. The stationary distribution is LogNormal(0.8, 0.5) and we applied an Exp(1) hyperprior on the relaxation parameter of the process. This prior constrains Re to a wide range of reasonable values (95% range 0.8 – 5.9) and penalizes large changes in Re from week-to-week. Finally, we allowed the sampling proportion to vary when Swiss testing or genome sampling regimes changed significantly (Table S3). For our main analysis, we applied a broad LogUniform(10^−4^, 1) prior on the sampling proportion, since we do not know how many individuals were truly infected. Alternatively, we also tried a LogUniform(10^−4^, 0.05) prior since we upper-bounded our sampling to 5% of confirmed cases each week (Supplementary text S3).

Finally, we added an additional transmission damping factor to the model. This factor is a multiplicative damping of Re applied to each introduction from 2 days after the oldest to the most-recent sampling date in the introduction. Since we hypothesized contact tracing was not functioning as well during periods of high case numbers, we estimated a separate damping factor for each of three periods: before 15 June 2020 (spring), 15 June to 30 September 2020 (summer), and 30 September to 1 December 2020 (fall). We used the same uninformative spike and slab prior for the damping factor in each period, with an inclusion probability of 0.5 and a uniform prior between 0 and 1, if included. For a description of the implementation of this model extension, see the Supplementary text S4.

For each phylodynamic model configuration (bounded and unbounded sampling proportion prior, with and without the contact tracing damping factor) and set of introductions (many and few), we ran five independent MCMC chains. We discarded the first 10% of each chain as burn-in and combined the remaining samples across the five chains. We evaluated the effective sample size (ESS) using Tracer (*51*) and verified that the ESS was at least 100 for all inferred parameters.

### New Zealand analysis

Genome sequence selection was done as for the Swiss analysis, except that we down-sampled available sequences from GISAID to 40% of confirmed case counts each week rather than 5% and we used national case count numbers rather than stratified by region. Phylogenetic analysis was performed as for the Swiss data. The phylodynamic analysis was also the same, except that we assumed a constant sampling proportion through time and for the bounded sampling proportion prior we used a LogUniform(10^−4^, 0.4) prior to match the down-sampling scheme.

## Data Availability

All genome sequence data used in the analysis is available on GISAID (https://www.gisaid.org/). Data generated by the Swiss SARS-CoV-2 Sequencing Consortium is available on GISAID (submitting lab: Department of Biosystems Science and Engineering, ETH Zuerich). The code used to generate figures and values for the manuscript is available at https://github.com/SarahNadeau/cov-swiss-phylo. The phylogenetic analysis code is at https://github.com/cevo-public/Grapevine-SARS-CoV-2-Introduction-Analysis and the phylodynamic analysis code, including BEAST2 XML files, is at https://github.com/tgvaughan/TransmissionChainAnalyses.

## List of Supplementary Materials

Supplementary text S1 to S4

Fig S1 to S10

Tables S1 to S3

## Acknowledgments

We gratefully acknowledge the authors from the originating laboratories responsible for obtaining the specimens and the submitting laboratories where genetic sequence data were generated and shared via the GISAID Initiative, on which this research is based. Joep de Ligt, David Winter, and Jing Wang kindly provided additional information on the source of analyzed New Zealand sequences. A full acknowledgements table of the contributing groups, including the identifiers for all GISAID data used in this study, is available on the project GitHub repository at https://github.com/SarahNadeau/cov-swiss-phylo. We thank Jana Huisman for valuable discussions on the manuscript.

## Funding

This work was supported by:

Swiss National Science Foundation grant 31CA30_196267 (TS)

ETH Zurich

## Author contributions

Conceptualization: SN, TV, TS

Data curation: SN, CB, IT, CC, TS, IN, NS, EB, PF, KPJ, SPC, SS, NSS, JMMG, PC, PPB, MPL, VK, SS, MZ, MH, AT, SC, FL, ARG, SA, TP, DJ, CB, GG, KL, MS, AM, TR, HSS, HHH, AE, NB, CB

Formal Analysis: SN, TV

Funding acquisition: TS

Methodology: SN, TV, LDP, EH, RAN, TS

Project administration: TS

Resources: VC, MR, OK, CN

Software: TV, CC Supervision: TS

Visualization: SN, TV

Writing – original draft: SN, TS

Writing – review & editing: SN, TV, CB, IT, CC, EH, TS, IN, NS, EB, PF, KPJ, SPC, CV, SS, NSS, JMMG, PC, PPB, MPL, VK, SS, MZ, MH, AT, SC, FL, ARG, SA, TP, DJ, CB, GG, KL, MS, AM, TR, HSS, HHH, AE, MR, OK, CN, LDP, NB, RAN, CB, TS

## Data and materials availability

All genome sequence data used in the analysis is available on GISAID (gisaid.org). Data generated by the Swiss SARS-CoV-2 Sequencing Consortium is available on GISAID (submitting lab: Department of Biosystems Science and Engineering, ETH Zürich). The code used to generate figures and values for the manuscript is available at https://github.com/SarahNadeau/cov-swiss-phylo. The phylogenetic analysis code is at https://github.com/cevo-public/Grapevine-SARS-CoV-2-Introduction-Analysis and the phylodynamic analysis code, including BEAST2 XML files, is at https://github.com/tgvaughan/TransmissionChainAnalyses.

## References and Notes

1. B. B. Oude Munnink, N. Worp, D. F. Nieuwenhuijse, R. S. Sikkema, B. Haagmans, R. A. M. Fouchier, M. Koopmans, The next phase of SARS-CoV-2 surveillance: real-time molecular epidemiology. Nat. Med. 27, 1518–1524 (2021).

2. M. U. G. Kraemer, D. A. T. Cummings, S. Funk, R. C. Reiner, N. R. Faria, O. G. Pybus, S. Cauchemez, Reconstruction and prediction of viral disease epidemics. Epidemiol. Infect. 147, e34 (2018).

3. R. J. Rockett, A. Arnott, C. Lam, R. Sadsad, V. Timms, K.-A. Gray, J.-S. Eden, S. Chang, M. Gall, J. Draper, E. M. Sim, N. L. Bachmann, I. Carter, K. Basile, R. Byun, M. V. O’Sullivan, S. C.-A. Chen, S. Maddocks, T. C. Sorrell, D. E. Dwyer, E. C. Holmes, J. Kok, M. Prokopenko, V. Sintchenko, Revealing COVID-19 transmission in Australia by SARS-CoV-2 genome sequencing and agent-based modeling. Nat. Med. 26, 1398–1404 (2020).

4. J. Douglas, F. K. Mendes, R. Bouckaert, D. Xie, C. L. Jiménez-Silva, C. Swanepoel, J. de Ligt, X. Ren, M. Storey, J. Hadfield, C. R. Simpson, J. L. Geoghegan, A. J. Drummond, D. Welch, Phylodynamics reveals the role of human travel and contact tracing in controlling the first wave of COVID-19 in four island nations. Virus Evol. 7, veab052 (2021).

5. P. W. G. Mallon, F. Crispie, G. Gonzalez, W. Tinago, A. A. Garcia Leon, M. McCabe, E. de Barra, O. Yousif, J. S. Lambert, C. J. Walsh, J. G. Kenny, E. Feeney, M. Carr, P. Doran, P. D. Cotter, Whole-genome sequencing of SARS-CoV-2 in the Republic of Ireland during waves 1 and 2 of the pandemic. medRxiv (2021),, doi:10.1101/2021.02.09.21251402.

6. L. du Plessis, J. T. McCrone, A. E. Zarebski, V. Hill, C. Ruis, B. Gutierrez, J. Raghwani, J. Ashworth, R. Colquhoun, T. R. Connor, N. R. Faria, B. Jackson, N. J. Loman, Á. O’Toole, S. M. Nicholls, K. V. Parag, E. Scher, T. I. Vasylyeva, E. M. Volz, A. Watts, I. I. Bogoch, K. Khan, COVID-19 Genomics UK (COG-UK) Consortium, D. M. Aanensen, M. U. G. Kraemer, A. Rambaut, O. G. Pybus, Establishment and lineage dynamics of the SARS-CoV-2 epidemic in the UK. Science. 371, 708–712 (2021).

7. M. Stange, A. Mari, T. Roloff, H. M. Seth-Smith, M. Schweitzer, M. Brunner, K. Leuzinger, K. K. Søgaard, A. Gensch, S. Tschudin-Sutter, S. Fuchs, J. Bielicki, H. Pargger, M. Siegemund, C. H. Nickel, R. Bingisser, M. Osthoff, S. Bassetti, R. Schneider-Sliwa, M. Battegay, H. H. Hirsch, A. Egli, SARS-CoV-2 outbreak in a tri-national urban area is dominated by a B.1 lineage variant linked to a mass gathering event. PLoS Pathog. 17, e1009374 (2021).

8. D. Miller, M. A. Martin, N. Harel, O. Tirosh, T. Kustin, M. Meir, N. Sorek, S. Gefen-Halevi, S. Amit, O. Vorontsov, A. Shaag, D. Wolf, A. Peretz, Y. Shemer-Avni, D. Roif-Kaminsky, N. M. Kopelman, A. Huppert, K. Koelle, A. Stern, Full genome viral sequences inform patterns of SARS-CoV-2 spread into and within Israel. Nat. Commun. 11, 5518 (2020).

9. J. L. Geoghegan, X. Ren, M. Storey, J. Hadfield, L. Jelley, S. Jefferies, J. Sherwood, S. Paine, S. Huang, J. Douglas, F. K. Mendes, A. Sporle, M. G. Baker, D. R. Murdoch, N. French, C. R. Simpson, D. Welch, A. J. Drummond, E. C. Holmes, S. Duchêne, J. de Ligt, Genomic epidemiology reveals transmission patterns and dynamics of SARS-CoV-2 in Aotearoa New Zealand. Nat. Commun. 11, 6351 (2020).

10. N. F. Müller, C. Wagner, C. D. Frazar, P. Roychoudhury, J. Lee, L. H. Moncla, B. Pelle, M. Richardson, E. Ryke, H. Xie, L. Shrestha, A. Addetia, V. M. Rachleff, N. A. P. Lieberman, M.-L. Huang, R. Gautom, G. Melly, B. Hiatt, P. Dykema, A. Adler, E. Brandstetter, P. D. Han, K. Fay, M. Ilcisin, K. Lacombe, T. R. Sibley, M. Truong, C. R. Wolf, M. Boeckh, J. A. Englund, M. Famulare, B. R. Lutz, M. J. Rieder, M. Thompson, J. S. Duchin, L. M. Starita, H. Y. Chu, J. Shendure, K. R. Jerome, S. Lindquist, A. L. Greninger, D. A. Nickerson, T. Bedford, Viral genomes reveal patterns of the SARS-CoV-2 outbreak in Washington State. Sci. Transl. Med. 13 (2021), doi:10.1126/scitranslmed.abf0202.

11. S3C, Swiss SARS-CoV-2 Sequencing Consortium (S3C) (2020), (available at https://web.archive.org/web/20211021102940/https://bsse.ethz.ch/cevo/research/sars-cov-2/swiss-sars-cov-2-sequencing-consortium.html).

12. P. Bogner, I. Capua, D. J. Lipman, N. J. Cox, A global initiative on sharing avian flu data. Nature Publishing Group UK (2006),, doi:10.1038/442981a.

13. World Health Organization, Tracking SARS-CoV-2 variants (2021), (available at https://web.archive.org/web/20211104163350/https://www.who.int/en/activities/tracking-SARS-CoV-2-variants/).

14. A. Rambaut, E. C. Holmes, Á. O’Toole, V. Hill, J. T. McCrone, C. Ruis, L. du Plessis, O. G. Pybus, A dynamic nomenclature proposal for SARS-CoV-2 lineages to assist genomic epidemiology. Nat Microbiol. 5, 1403–1407 (2020).

15. S. Bradley, Switzerland re-opens its European borders. swissinfo.ch (2020), (available at https://web.archive.org/web/20211020095340/https://www.swissinfo.ch/eng/politics/covid-19_what-s-happening-at-swiss-borders-and-airports-/45727184).

16. T. Stadler, D. Kühnert, S. Bonhoeffer, A. J. Drummond, Birth-death skyline plot reveals temporal changes of epidemic spread in HIV and hepatitis C virus (HCV). Proc. Natl. Acad. Sci. U. S. A. 110, 228–233 (2013).

17. Swiss Federal Statistical Office, Federal Statistical Office (2020), (available at https://www.bfs.admin.ch/bfs/en/home.html).

18. T. Stadler, Sampling-through-time in birth-death trees. J. Theor. Biol. 267, 396–404 (2010).

19. Swiss Federal Office of Public Health, COVID-19 Switzerland (2021), (available at https://web.archive.org/web/20211104165544/https://www.covid19.admin.ch/en/epidemiologic/case).

20. B. Marquis, O. Opota, K. Jaton, G. Greub, Impact of different SARS-CoV-2 assays on laboratory turnaround time. J. Med. Microbiol. 70 (2021), doi:10.1099/jmm.0.001280.

21. B. Morel, P. Barbera, L. Czech, B. Bettisworth, L. Hübner, S. Lutteropp, D. Serdari, E.-G. Kostaki, Mamais, A. M. Kozlov, P. Pavlidis, D. Paraskevis, A. Stamatakis, Phylogenetic analysis of SARS-CoV-2 data is difficult. bioRxiv (2020),, doi:10.1101/2020.08.05.239046.

22. European Centre for Disease Prevention and Control, Download historical data (to 14 December 2020) on the daily number of new reported COVID-19 cases and deaths worldwide. European Centre for Disease Prevention and Control (2020), (available at https://www.ecdc.europa.eu/en/publications-data/download-todays-data-geographic-distribution-covid-19-cases-worldwide).

23. New Zealand Government, History of the COVID-19 Alert System. Unite against COVID-19 (2021), (available at https://web.archive.org/web/20211020111429/https://covid19.govt.nz/alert-levels-and-updates/history-of-the-covid-19-alert-system/).

24. J. L. Geoghegan, J. Douglas, X. Ren, M. Storey, J. Hadfield, O. K. Silander, N. E. Freed, L. Jelley, S. Jefferies, J. Sherwood, S. Paine, S. Huang, A. Sporle, M. G. Baker, D. R. Murdoch, A. J. Drummond, D. Welch, C. R. Simpson, N. French, E. C. Holmes, J. de Ligt, Use of Genomics to Track Coronavirus Disease Outbreaks, New Zealand. Emerg. Infect. Dis. 27, 1317–1322 (2021).

25. The Swiss Federal Council, Federal Council to gradually ease measures against the new coronavirus (2020), (available at https://web.archive.org/web/20220527142858/https://www.admin.ch/gov/en/start/documentation/media-releases.msg-id-78818.html).

26. A. Ladoy, O. Opota, P.-N. Carron, I. Guessous, S. Vuilleumier, S. Joost, G. Greub, Size and duration of COVID-19 clusters go along with a high SARS-CoV-2 viral load: A spatio-temporal investigation in Vaud state, Switzerland. Sci. Total Environ. 787, 147483 (2021).

27. J. C. Lemaitre, J. Perez-Saez, A. S. Azman, A. Rinaldo, J. Fellay, Assessing the impact of non-pharmaceutical interventions on SARS-CoV-2 transmission in Switzerland. Swiss Med. Wkly. 150, w20295 (2020).

28. J. S. Huisman, J. Scire, D. C. Angst, J. Li, R. A. Neher, M. H. Maathuis, S. Bonhoeffer, T. Stadler, Estimation and worldwide monitoring of the effective reproductive number of SARS-CoV-2. medRxiv (2021),, doi:10.1101/2020.11.26.20239368.

29. E. B. Hodcroft, M. Zuber, S. Nadeau, T. G. Vaughan, K. H. D. Crawford, C. L. Althaus, M. L. Reichmuth, J. E. Bowen, A. C. Walls, D. Corti, J. D. Bloom, D. Veesler, D. Mateo, A. Hernando, I. Comas, F. González-Candelas, SeqCOVID-SPAIN consortium, T. Stadler, R. A. Neher, Spread of a SARS-CoV-2 variant through Europe in the summer of 2020. Nature. 595, 707–712 (2021).

30. Keystone-SDA, Contact tracing not working properly, writes paper. swissinfo.ch (2020), (available at https://web.archive.org/web/20211020105928/https://www.swissinfo.ch/eng/business/contact-tracing-not-working-properly--writes-paper/46090060).

31. T. Fetzer, T. Graeber, Measuring the scientific effectiveness of contact tracing: Evidence from a natural experiment. Proc. Natl. Acad. Sci. U. S. A. 118 (2021), doi:10.1073/pnas.2100814118.

32. The ARTIC Network, artic-ncov2019/primer_schemes/nCoV-2019/V3 at master · artic-network/artic-ncov2019. GitHub (2020), (available at https://github.com/artic-network/artic-ncov2019).

33. S. Posada-Céspedes, D. Seifert, I. Topolsky, K. P. Jablonski, K. J. Metzner, N. Beerenwinkel, V-pipe: a computational pipeline for assessing viral genetic diversity from high-throughput data. Bioinformatics (2021), doi:10.1093/bioinformatics/btab015.

34. R. Schmieder, R. Edwards, Quality control and preprocessing of metagenomic datasets. Bioinformatics. 27, 863–864 (2011).

35. F. Wu, S. Zhao, B. Yu, Y.-M. Chen, W. Wang, Z.-G. Song, Y. Hu, Z.-W. Tao, J.-H. Tian, Y.-Y. Pei, M.-L. Yuan, Y.-L. Zhang, F.-H. Dai, Y. Liu, Q.-M. Wang, J.-J. Zheng, L. Xu, E. C. Holmes, Y.-Z. Zhang, A new coronavirus associated with human respiratory disease in China. Nature. 579, 265–269 (2020).

36. H. Li, R. Durbin, Fast and accurate short read alignment with Burrows-Wheeler transform. Bioinformatics. 25, 1754–1760 (2009).

37. I. Aksamentov, C. Roemer, E. Hodcroft, R. Neher, Nextclade: clade assignment, mutation calling and quality control for viral genomes. J. Open Source Softw. 6, 3773 (2021).

38. K. Katoh, K. Misawa, K.-I. Kuma, T. Miyata, MAFFT: a novel method for rapid multiple sequence alignment based on fast Fourier transform. Nucleic Acids Res. 30, 3059–3066 (2002).

39. Nextstrain, nextstrain/ncov. GitHub (2020), (available at https://github.com/nextstrain/ncov).

40. N. De Maio, C. Walker, R. Borges, L. Weilguny, G. Slodkowicz, N. Goldman, Issues with SARS-CoV-2 sequencing data. Virological (2020), (available at https://virological.org/t/issues-with-sars-cov-2-sequencing-data/473).

41. L.-T. Nguyen, H. A. Schmidt, A. von Haeseler, B. Q. Minh, IQ-TREE: a fast and effective stochastic algorithm for estimating maximum-likelihood phylogenies. Mol. Biol. Evol. 32, 268–274 (2015).

42. M. Hasegawa, H. Kishino, T. Yano, Dating of the human-ape splitting by a molecular clock of mitochondrial DNA. J. Mol. Evol. 22, 160–174 (1985).

43. Z. Yang, Maximum likelihood phylogenetic estimation from DNA sequences with variable rates over sites: approximate methods. J. Mol. Evol. 39, 306–314 (1994).

44. T.-H. To, M. Jung, S. Lycett, O. Gascuel, Fast Dating Using Least-Squares Criteria and Algorithms. Syst. Biol. 65, 82–97 (2016).

45. Nextstrain, Genomic epidemiology of SARS-CoV-2 with subsampling focused globally over the past 6 months. Nextstrain (2020), (available at https://nextstrain.org/ncov/gisaid/global/6m?l=clock).

46. K. Miyakawa, H. Narushima, Lattice-theoretic properties of MPR-posets in phylogeny. Discrete Appl. Math. 134, 169–192 (2004).

47. Swiss Federal Office of Public Health, Coronavirus: Frequently Asked Questions (FAQs) (2022), (available at https://web.archive.org/web/20220527153158/https://www.bag.admin.ch/bag/en/home/krankheiten/ausbrueche-epidemien-pandemien/aktuelle-ausbrueche-epidemien/novel-cov/haeufig-gestellte-fragen.html).

48. Schweizer Tourismus in Zahlen. STV-FST, (available at https://www.stv-fst.ch/de/stiz).

49. R. Bouckaert, T. G. Vaughan, J. Barido-Sottani, S. Duchêne, M. Fourment, A. Gavryushkina, J. Heled, G. Jones, D. Kühnert, N. De Maio, M. Matschiner, F. K. Mendes, N. F. Müller, H. A. Ogilvie, L. du Plessis, A. Popinga, A. Rambaut, D. Rasmussen, I. Siveroni, M. A. Suchard, C.-H. Wu, D. Xie, C. Zhang, T. Stadler, A. J. Drummond, BEAST 2.5: An advanced software platform for Bayesian evolutionary analysis. PLoS Comput. Biol. 15, e1006650 (2019).

50. Keystone-SDA, Switzerland confirms first coronavirus case. swissinfo.ch (2020), (available at https://web.archive.org/web/20220525122732/https://www.swissinfo.ch/eng/politics/covid-19_switzerland-confirms-first-coronavirus-case/45579278).

51. A. Rambaut, A. J. Drummond, D. Xie, G. Baele, M. A. Suchard, Posterior Summarization in Bayesian Phylogenetics Using Tracer 1.7. Syst. Biol. 67, 901–904 (2018).

