## Supplementary information for "Swiss public health measures associated with reduced SARS-CoV-2 transmission using genome data"

### Supplementary text

#### S1: Sensitivity analyses for identifying introductions

Here we describe different sensitivity analyses we performed for the definition of an introduction.

*Criteria for identifying introductions.* First, we assessed the sensitivity of identified introductions to the precise heuristic definition of an introduction. We began with the same lineage-specific phylogenetic trees generated for the main analysis. Then, we re-identified introductions from these trees while varying (a) the maximum number of export events allowed from each introduction and (b) the maximum number of consecutive export events allowed to occur along each single internal branch. Figure S2A shows that increasing (a) yields fewer, larger introductions. Increasing (b) for each level of (a) has a negligible effect. However, the greatest differences come from the different assumptions about how to resolve polytomies before applying these heuristics. Increasing (a) from 1 to 4 yields approximately 25% fewer introductions, while resolving polytomies such that Swiss descendants cluster together yields approximately 75% fewer introductions (Figure S2A). We chose to present results using an introduction definition based on (a) a maximum of three exported lineages and (b) a maximum one consecutive export on each internal branch. This allows for some exports from Swiss introductions but not arbitrarily many. We rely on our different polytomy assumptions to capture most of the uncertainty in the number and size of introductions.

*Ratio of foreign context to focal Swiss sequences analyzed.* Next, we assessed the sensitivity of identified introductions to the sequence set analyzed. We re-sampled sequences to analyze three times, each time taking a number of Swiss sequences corresponding to up to 5% of confirmed cases each week. Then we sampled foreign context sequences at a 1:1, 2:1, and 3:1 ratio to the Swiss sequences. Figure S2B shows that as we add foreign context sequences, we identify more numerous, smaller introductions. However, the greatest differences come from the different assumptions about how to resolve polytomies (few vs. many introductions), not the ratio of foreign context to focal Swiss sequences. Therefore, we chose to present results using the 2:1 ratio to balance speed (smaller dataset = faster tree search convergence) and information content (larger dataset = more introductions represented).

*Number of focal Swiss sequences analyzed.* Finally, we assessed whether the number of identified introductions saturates as we add more Swiss sequences. To do this, we sub-sampled the Swiss genome sequences used in our main analysis to 20, 40, 60, and 80% of the full set of sequences analyzed, pruning the not-included Swiss sequences from the phylogenetic trees generated for the main analysis. Then, we calculated the number of introductions we would have identified on the pruned trees. We performed the random sub-sampling and pruning 50 times for each sub-sampling level. Figure S2C shows that as we approach the number of Swiss sequences used in the main analysis, we do not reach saturation. Therefore, if we were to include even more sequences, we would identify more introductions.

### S2: New Zealand validation data

For New Zealand, the sequence submitters provided additional information on which samples were from cases in managed isolation and quarantine (MIQ) facilities versus the broader community. This allows us to partially evaluate our introduction identification methods. 117 of the 1234 analyzed focal sequences in the New Zealand analysis originated from MIQ facilities. 63 (54%) of these were singletons under the "many introductions" polytomy assumption versus 37 (32%) under the "few introductions" assumption. 44 (38%) or 37 (32%) were plausible within-MIQ outbreaks. These were identified as introductions with cases all from a single region and all MIQ. They may represent groups of individuals quarantining together or infected in the same source location. These outbreaks included, on average, 3 samples spanning 5 days (many introductions) or 2 samples spanning 9 days (few introductions). The remaining 10 (9%) or 43 (37%) of MIQ sequences were in introductions including community cases or including cases in multiple MIQ facilities in different regions, which we deem unrealistic. These results support that the "many introductions" polytomy assumption is more realistic when the probability of infection abroad is high compared to the probability of locally acquired infection.

### S3: Sensitivity analyses for phylodynamic modeling

Here we describe a sensitivity analysis and some example intermediate outputs from our phylodynamic analysis.

*Sampling proportion prior*. We repeated our analyses using two different priors on the sampling proportion. The first, unbounded prior was LogUniform($10^{-4}$, 1). This broad prior allows the sampling proportion to assume any value. The second, bounded prior was LogUniform($10^{-4}$, 0.05). This narrower prior is motivated by our 5% down-sampling based on confirmed case numbers. Figure S7A shows that in Switzerland, the estimated sampling proportion in late fall 2020 varies greatly depending on the prior. The rise in prevalence of lineage B.1.177 during this period (*29*), representing a drop in SARS-CoV-2 diversity in Switzerland, might explain why the inference under the broader sampling prior estimates a proportion corresponding to fewer individuals than we know were infected during this time. Figure S7B shows that the effective reproductive number estimates in fall 2020 for Switzerland more closely match estimates based on confirmed case data when the sampling proportion is treated as a fitting parameter, i.e., under the first, broad prior. Therefore, we report results under this prior in the main text. In Figure S8A, we show that the damping factor results are qualitatively similar between the two sampling proportion priors. For the New Zealand analysis, Re estimates are not affected by bounding the sampling proportion or not (Figure S10).

*Logged trees.* Finally, we visually inspected phylogenetic trees for a few introductions. These trees were sampled and logged by the Markov chains in the phylodynamic analyses. Note that the damping factor results are jointly inferred from all the branching events across introductions in each time period. For each set of model assumptions and each month, we inspected maximum clade credibility summary trees for the 50th and 95th percentile largest introductions that were first sampled that month and eventually yielded >2 samples. Figure S9 shows as an example summary trees for these introductions from one of the MCMC chains in the phylodynamic analysis for Switzerland with damping factors and an unbounded sampling proportion prior.

### S4: BDSKY introduction correction and damping factor extension

First we describe the correction applied to the number of observed introductions each week based on the BDSKY model. Then we describe the implementation of an extension to the BDSKY model to incorporate a transmission damping factor.

*Correction to the number of observed introductions*. The correction described here is to account for the time-varying Re in Switzerland, which affects the probability a new introduction each week went un-sampled through the end of the sampling period. The equations are taken from Stadler et al. (2013).

Let the birth rate $\lambda$ change weekly at times $t_i$ $(i = 1, \dots, m-1)$, where the process ends at time $m$ (the end of the sampling period). The death rate $\mu$ and sampling rate $\varphi$ are assumed to be constant through time. Let $A$ and $B$ be convenience parameters to simplify notation. They are defined as follows:

$$A_i = \sqrt{(\lambda_i - \mu - \varphi)^2 + 4\lambda_i\varphi}$$

$$B_i = \frac{\left(1 - 2p_{i+1}(t_i)\right)\lambda_i + \mu + \varphi}{A_i}$$

where $p_i(t)$ is the probability that an infected individual at time $t$ has no sampled descendants when the process ends. It is defined as follows:

$$p_i(t) = \frac{\lambda_i + \mu + \varphi - A_i \dfrac{e^{A_i(t_i - t)}(1 + B_i) - (1 - B_i)}{e^{A_i(t_i - t)}(1 + B_i) + (1 - B_i)}}{2\lambda_i}$$

In pseudocode, $p_i(t)$ is calculated for each weekly change time $t_i$ as follows:

1. Calculate $A_i$ for each time period $i$.
2. Calculate $B_m$ using $p_{m+1}(t_m) = 1$ since the probability an infected individual at time $m$ having no sampled descendants is 1.
3. For $(i = m, \dots, 1)$, calculate first $p_i$ and then $B_{i-1}(p_i)$.

Finally, the number of observed introductions each week $N_{o,i}$ is corrected for the probability a new introduction at the start of that week went un-sampled through the end of the sampling period:

$$N_i = \frac{N_{o,i}}{1 - p_i}$$

*Implementation of the transmission damping factor.* This section describes the implementation of a "damping factor" extension to the parameterization of the BDSKY phylodynamic model presented in Stadler et al. 2013.

As described in the main text, the damping factor is a multiplicative factor applied to the birth rate two days after the first sample in each identified introduction was collected. The value of the damping factor applied to each introduction is allowed to change at two times (representing the start of summer and start of fall).

This parameter configuration is an input to the new BEAST2 parameterization "EpiParameterizationMod", available at https://github.com/tgvaughan/BDMM-Prime. The probability density of each sampled tree is identical to that presented in Stadler et al. 2013 where the piecewise-constant birth rate $\lambda_i$ of a tree is modulated by the appropriate damping factor beginning two days after the first sample.

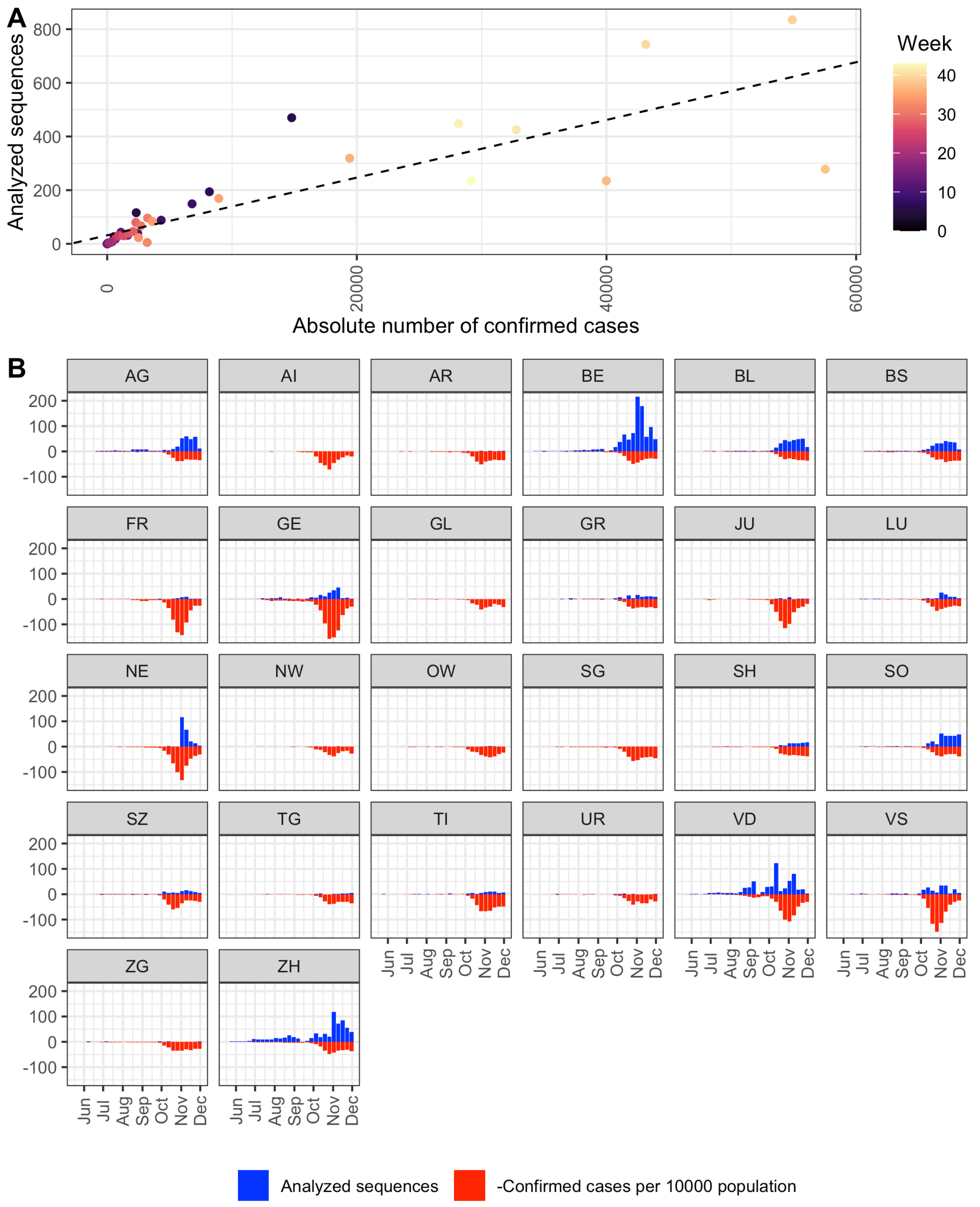


**Fig. S1.** Number of analyzed sequences compared to confirmed case counts each week for (**A**) all of Switzerland and (**B**) stratified by canton after the week of 18 May 2020, when case count data is also stratified by canton. The best-fit line in (A) has an $R^2$ value of 0.72. Week 0 corresponds to the start of sampling with the first sequence from Switzerland collected on 24 February 2020. Facet names in (B) are standard abbreviations for Swiss cantons.

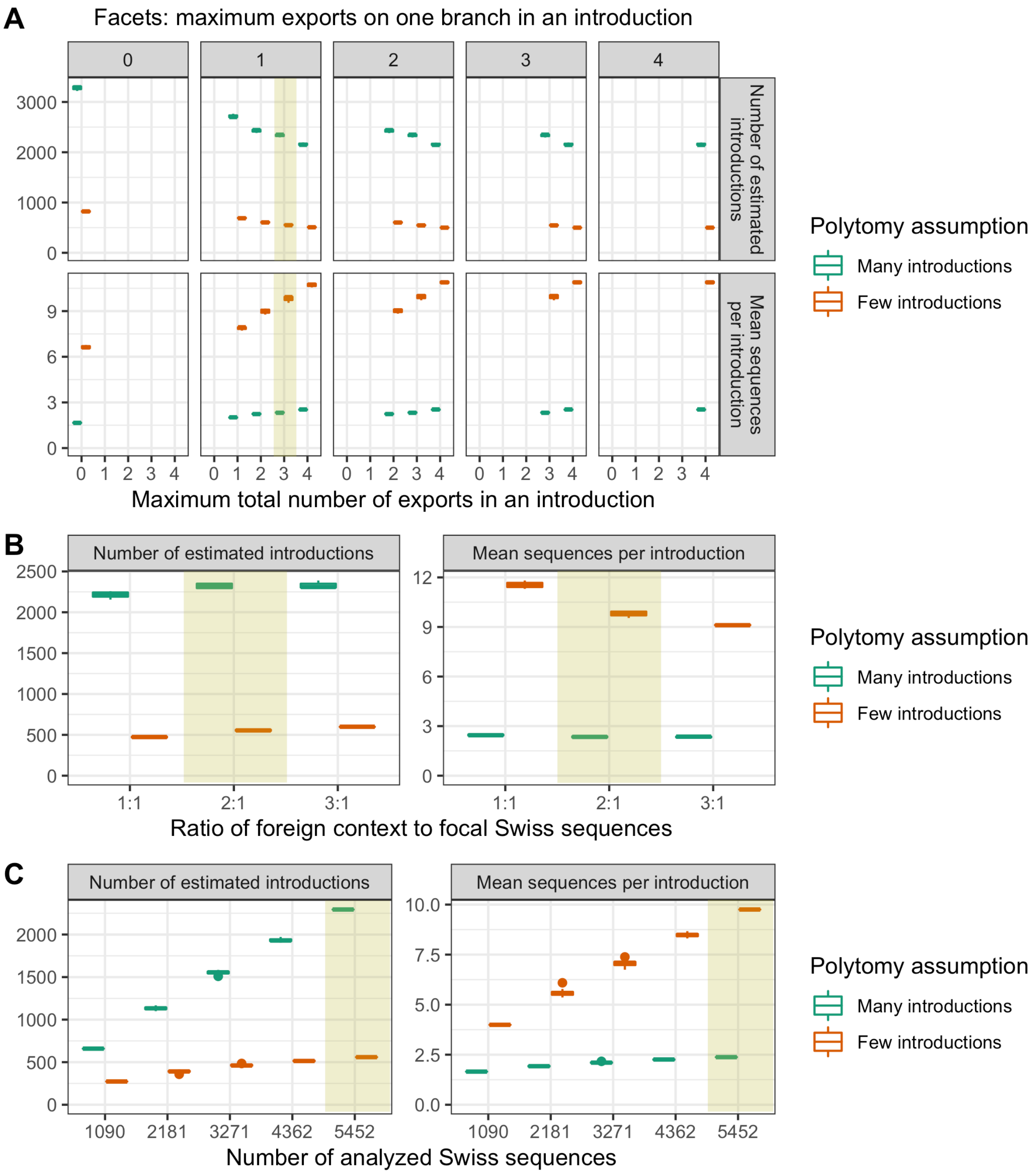


**Fig. S2.** Sensitivity analyses showing how summary statistics on the number and size of identified Swiss introductions change depending on the definition of an introduction. See Supplementary text S1 for details of the sensitivity analyses. (**A**) shows sensitivity to the heuristic thresholds used to define an introduction based on the lineage phylogenies, (**B**) shows sensitivity to the ratio of foreign context to focal sequences analyzed, and (**C**) shows sensitivity to the number of focal sequences analyzed. All statistics were generated under two different polytomy assumptions giving rise to either few or many introductions. Boxplots in (A) and (B) are for 3 randomly drawn datasets, boxplots in (C) are for 50 random sub-samples from the same dataset. Shaded yellow rectangles highlight values used for the main analysis.

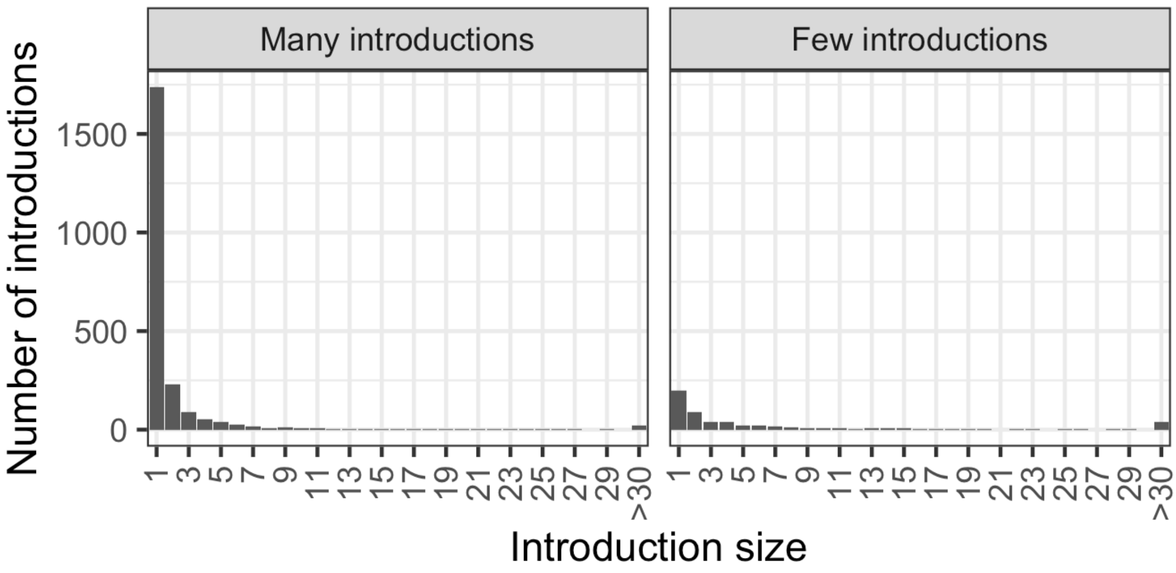


**Fig. S3.** Size distribution of estimated Swiss introductions.

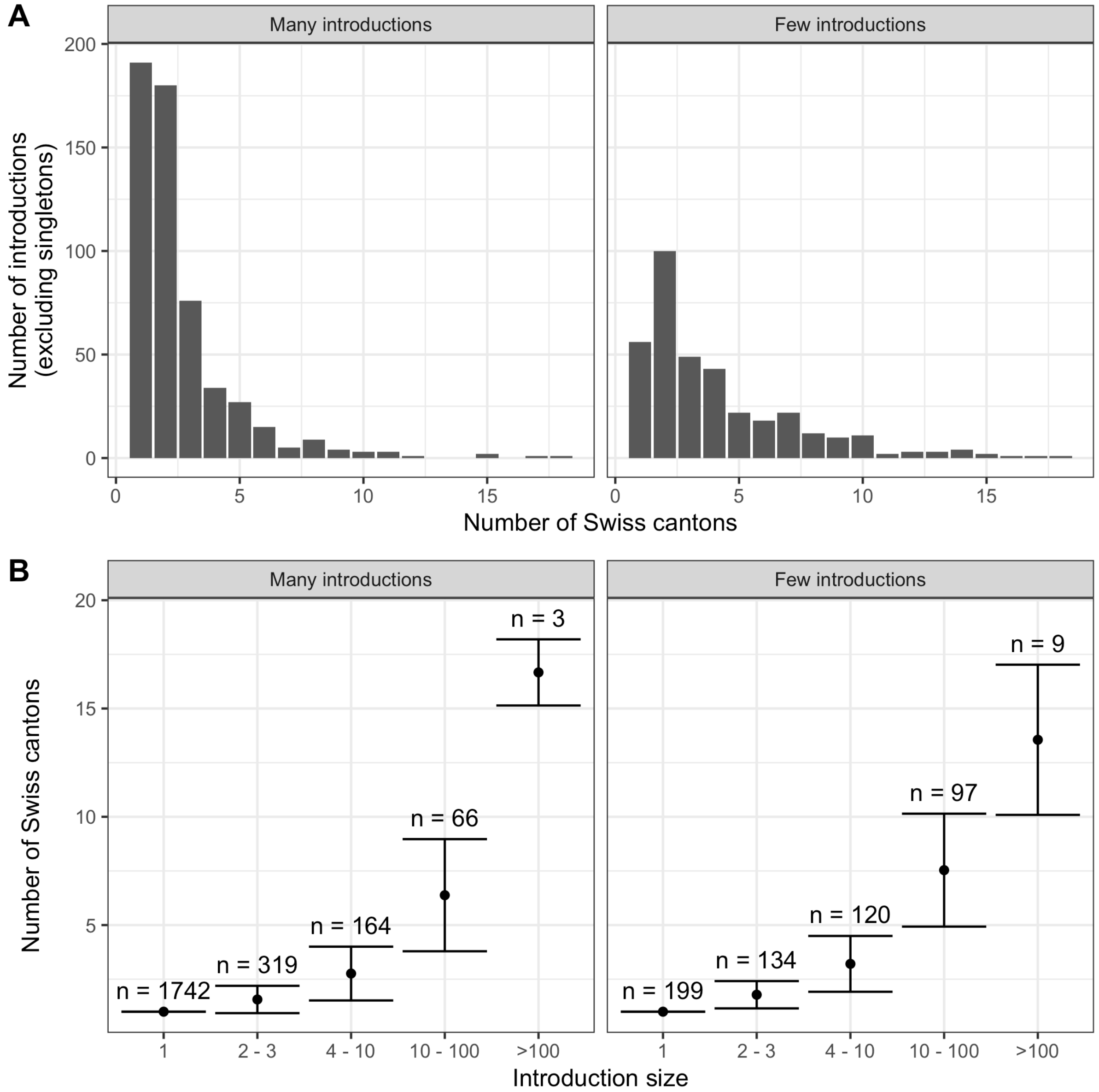


**Fig. S4.** Geographic distribution of Swiss introductions estimated under each polytomy assumption. (**A**) shows that most transmission chains were sampled in only one or two cantons. (**B**) shows that larger introductions were sampled in more Swiss cantons. Points show the mean number of cantons and error bars show the standard deviation in the number of cantons. Labels given the number of introductions in each size bin under each polytomy assumption.

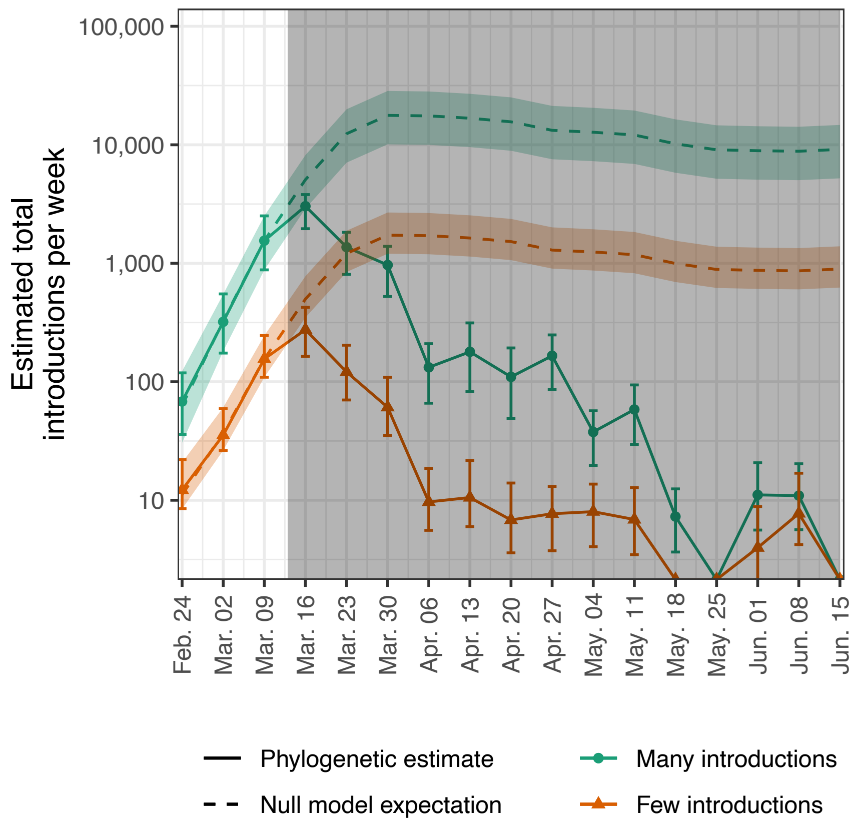


**Fig. S5.** Sensitivity analysis for modeling Swiss introductions. Estimated total introductions i.e., introductions scaled to account for time-varying sampling proportion (solid lines) are compared to a null model (dashed lines) where total introductions are a linear function of case numbers in all non-Swiss European countries, as defined in the European Centre for Disease Control (ECDC)'s case count data (*22*). The null model is fit to the points prior to the border closure on 13 March (highlighted with shaded rectangle), values after that are a model prediction. Uncertainty bounds for total introductions (error bars) and null model predictions (colored shaded areas) are based on the 95% upper and lower HPD bounds for Re when estimating total introductions. The orange and green colors correspond to estimates generated under our few and many introductions polytomy assumptions, respectively.

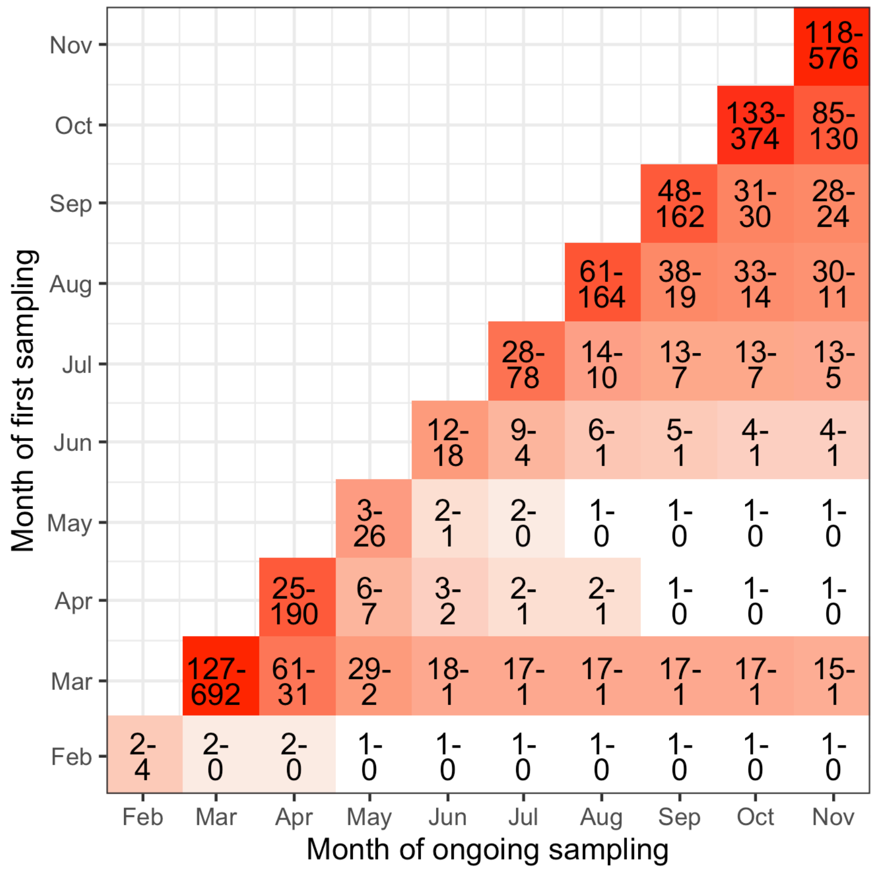


**Fig. S6.** Heatmap of the number of newly sampled introductions in Switzerland each month (diagonal entries) and the number continuing to persist into each following month (off-diagonal entries). Introductions are counted once in the month they are first sampled ("Month of first sampling") and one every following month ("Month of ongoing sampling") until the date of the latest sample. Estimates were generated under two different polytomy assumptions giving rise to either few or many introductions. Ranges are: few-many.

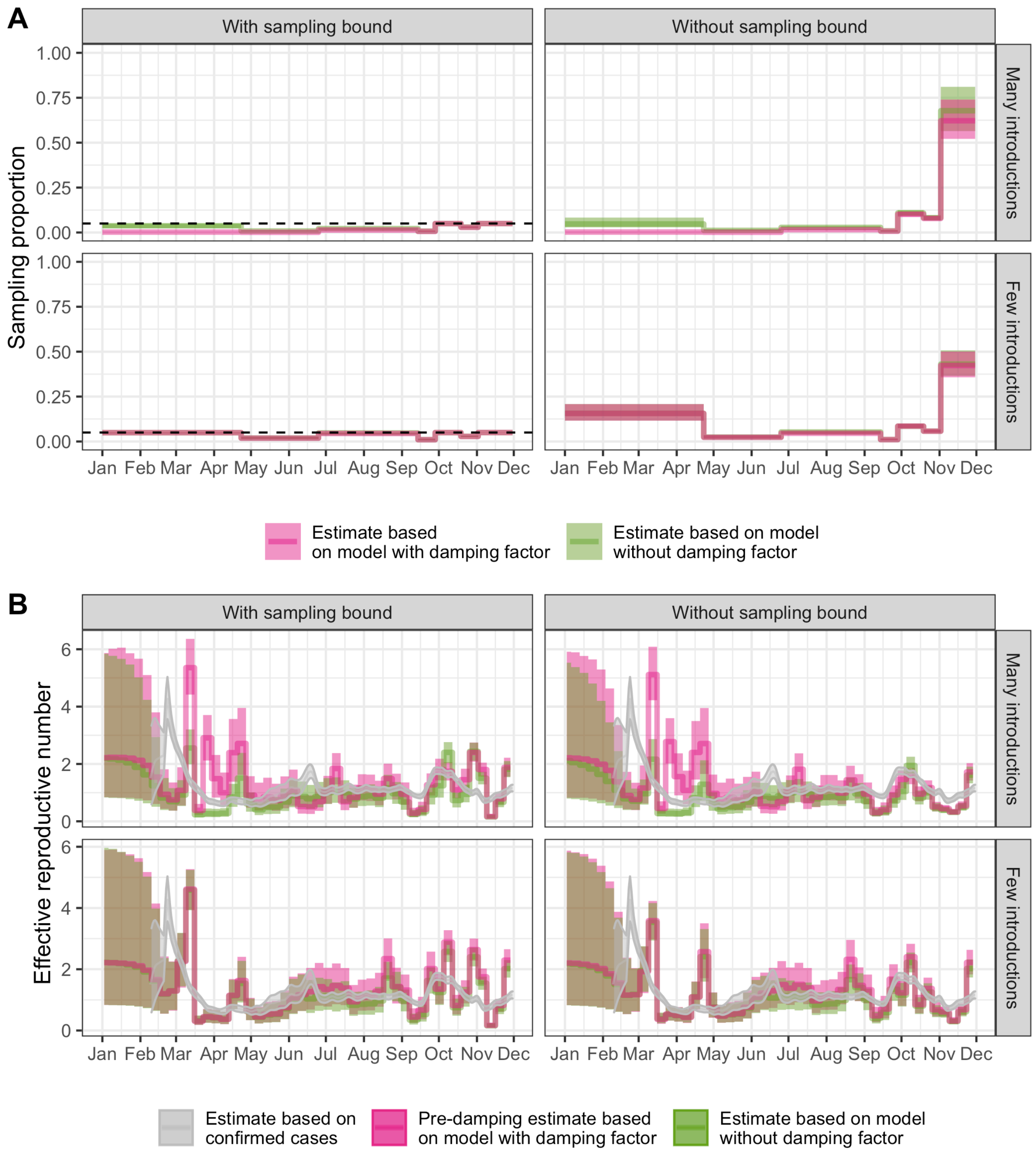


**Fig. S7.** Phylodynamic estimates for (**A**) the sampling proportion and (**B**) the time-varying effective reproductive number Re in Switzerland. The dashed line in (A) shows the sampling proportion prior's upper bound, if applicable. Re estimates in (B) are overlaid with estimates based on confirmed case count data (*28*) in gray. Additionally, Re estimates from the models with a damping factor (pink) are the "baseline" Re before introduction-specific damping (i.e., before application of a damping factor once introductions are older than 2-days post sampling).

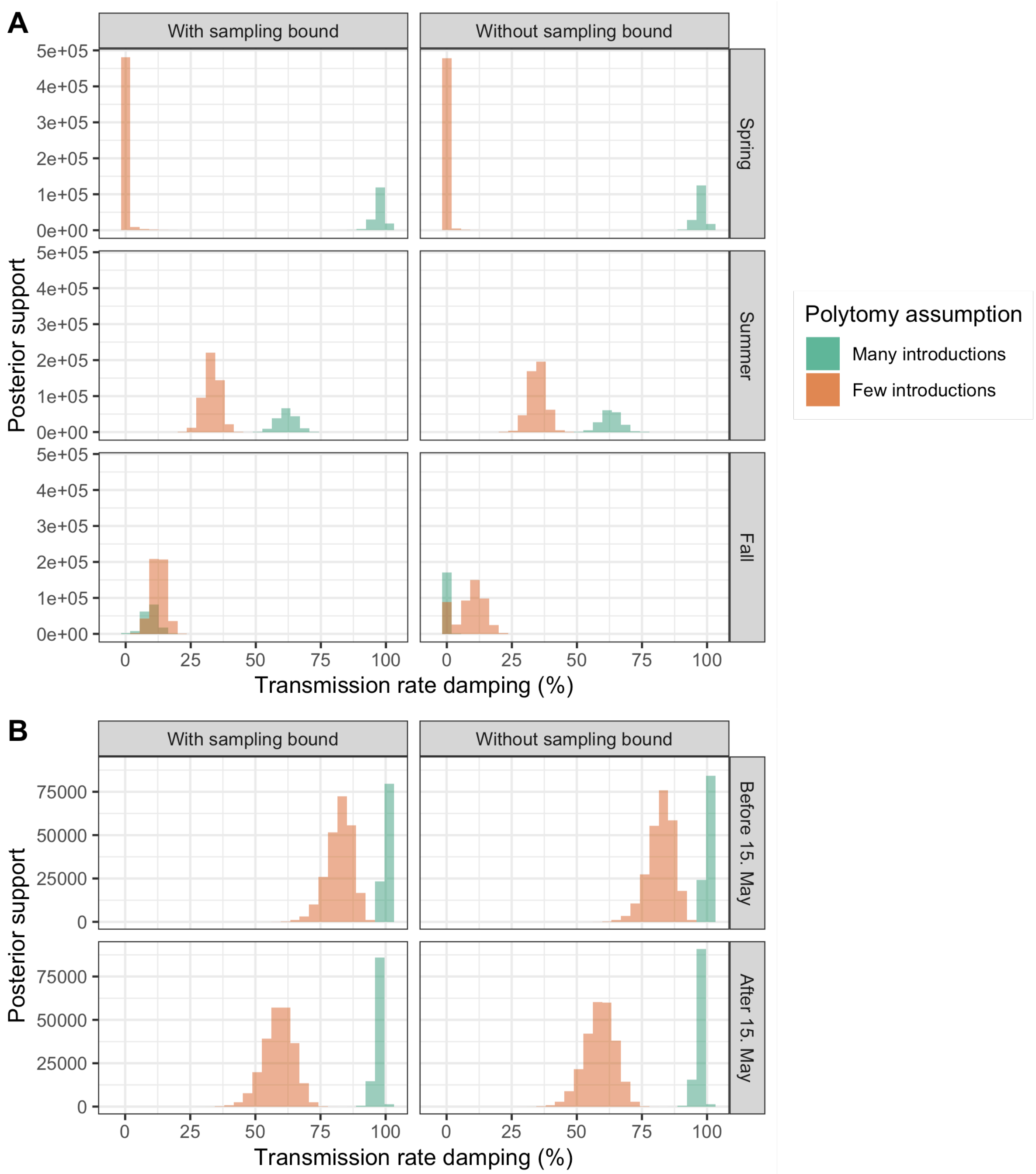


**Fig. S8.** Phylodynamic estimates for the damping factor in (**A**) Switzerland and (**B**) New Zealand in different time periods, conditioned on introductions estimated under two different polytomy assumptions giving rise to either few or many introductions.

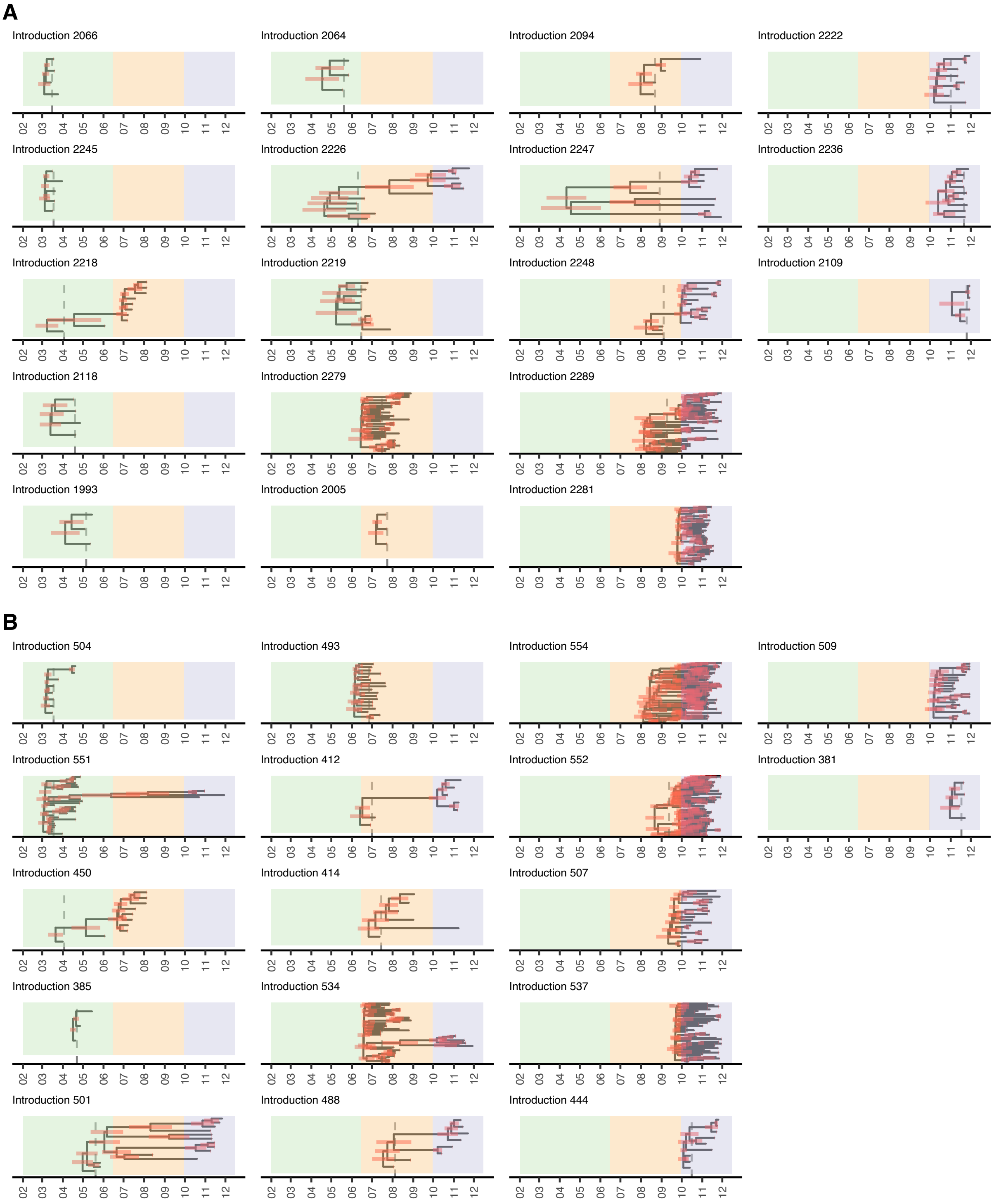


**Fig. S9.** A selection of maximum clade credibility summary trees from one of the MCMC chains in the phylodynamic analysis for Switzerland with damping factors and an unbounded sampling proportion prior. Here we show the 50th and 95th percentile introduction by size each month Mar – Nov 2020. Months are abbreviated by their number. (**A**) shows trees from the analysis conditioned on many introductions and (**B**) conditioned on few introductions. The three different color regions represent the spring (green), summer (orange) and fall (blue) periods. Vertical dashed lines show when the damping factor applies for each introduction - two days after the first sample date. Red bars show the 95% highest posterior density uncertainty in node dates.

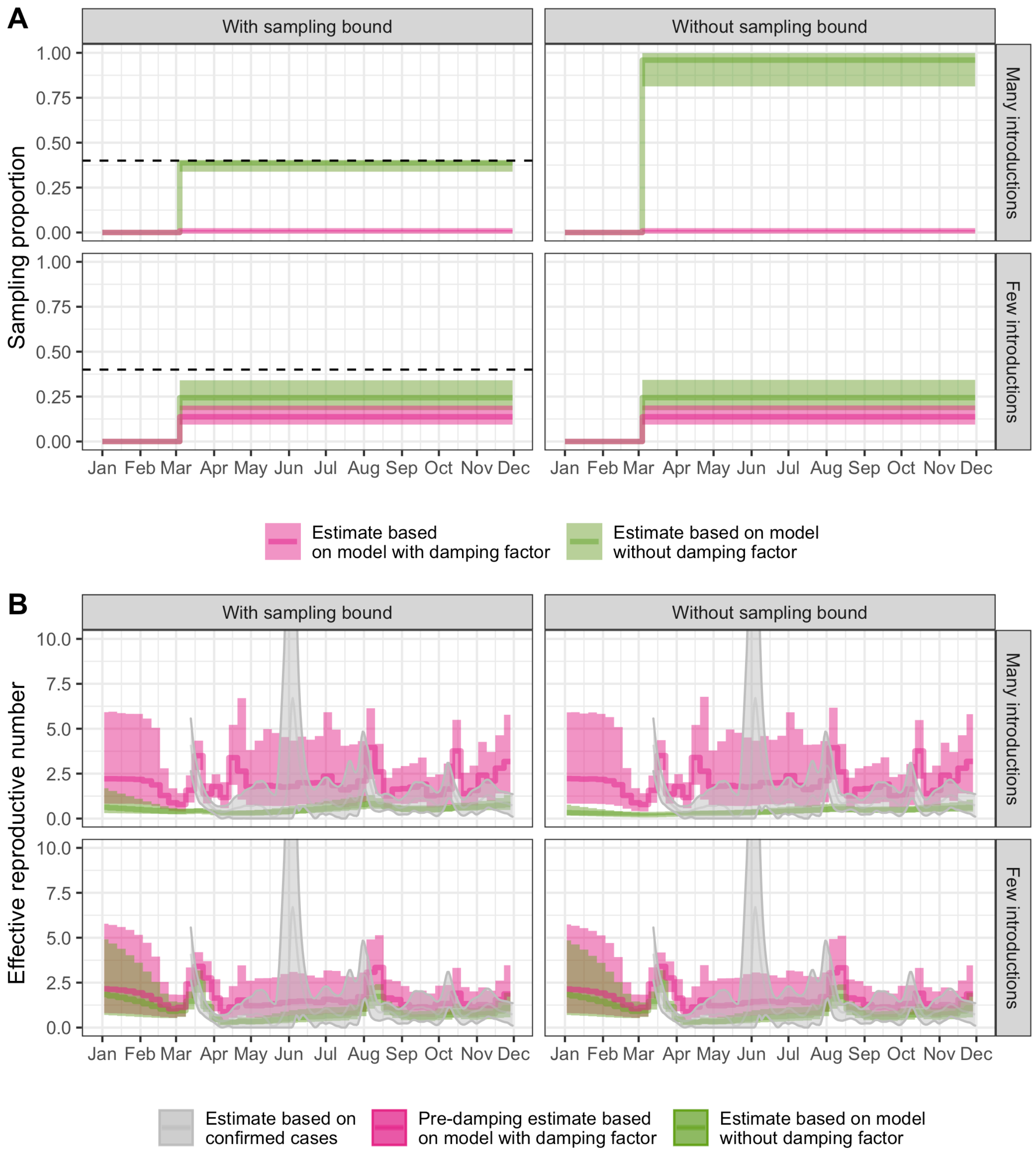


**Fig. S10.** Phylodynamic estimates for (**A**) the sampling proportion and (**B**) the time-varying effective reproductive number Re in New Zealand. The dashed line in (A) shows the sampling proportion prior's upper bound, if applicable. Re estimates in (B) are overlaid with estimates based on confirmed case count data (*28*) in gray. Additionally, Re estimates from the models with a damping factor (pink) are the "baseline" Re before introduction-specific damping (i.e. before application of a damping factor once introductions are older than 2-days post sampling).

**Table S1.** Summary of Pango lineages analyzed. If more than 50% of the samples from a lineage in the full, quality-filtered dataset were Swiss, we aggregated them into the parent lineage. The percentage of Swiss samples in the final, aggregated lineage sets are given in column "% lineage Swiss". Lineage aliases were also aggregated with their extended-form names. A separate phylogeny was constructed for each lineage analyzed.

| Lineage analyzed | Number Swiss samples analyzed | Lineages aggregated | % lineage Swiss |
|---|---|---|---|
| B.1.160 | 1347 | B.1.160, B.1.160.10, B.1.160.11, B.1.160.12, B.1.160.14, B.1.160.15, B.1.160.16, AB, B.1.160.19, B.1.160.20, B.1.160.22, B.1.160.26, B.1.160.29, B.1.160.30, B.1.160.31, B.1.160.32, B.1.160.9, B.1.160.16.1, AB.1 | 19 |
| B.1.177 | 1260 | B.1.177, B.1.177.23, B.1.177.28, B.1.177.43, B.1.177.44, B.1.177.71 | 4.8 |
| B.1 | 930 | B.1, B.1.214.2 | 2.2 |
| B.1.1 | 655 | B.1.1, B.1.1.144, B.1.1.327, B.1.1.39, AQ, B.1.1.524 | 3.1 |
| B.1.221 | 176 | B.1.221 | 8.1 |
| B.1.1.70 | 108 | B.1.1.70, AP | 15 |
| B.1.416.1 | 105 | B.1.416.1 | 45 |
| B.1.258 | 101 | B.1.258 | 4.4 |
| B.1.367 | 60 | B.1.367 | 10 |
| B.1.236 | 59 | B.1.236 | 33 |
| B.1.1.1.35 | 53 | B.1.1.1.35, C.35 | 13 |
| B.1.36.1 | 47 | B.1.36.1 | 35 |
| B.1.128 | 31 | B.1.128 | 3.3 |
| B.1.93 | 31 | B.1.93 | 3.3 |
| B.1.1.277 | 27 | B.1.1.277, K | 5.8 |
| B.1.1.47 | 24 | B.1.1.47 | 32 |
| B.1.1.269 | 19 | B.1.1.269 | 5 |
| B.1.1.1.36 | 16 | B.1.1.1.36, C.36, B.1.1.1.36.2, C.36.2 | 9.3 |
| B.1.1.10 | 16 | B.1.1.10, L | 2.7 |
| B.1.1.7 | 16 | B.1.1.7, Q | 0.85 |
| B.1.1.1 | 15 | B.1.1.1, C, B.1.1.1.5, C.5 | 0.86 |
| B.1.1.189 | 15 | B.1.1.189 | 12 |
| B.1.146 | 15 | B.1.146 | 30 |
| B.1.1.232 | 14 | B.1.1.232, AK | 3.1 |
| B | 11 | B | 0.44 |
| B.1.1.153 | 11 | B.1.1.153 | 6.5 |
| B.1.1.305 | 11 | B.1.1.305, AF, B.1.1.305.1, AF.1 | 8.4 |
| B.1.1.372 | 11 | B.1.1.372 | 0.95 |
| B.1.177.75 | 11 | B.1.177.75 | 12 |

| | | | |
|---|---|---|---|
| B.1.177.77 | 11 | B.1.177.77 | 6.1 |
| B.1.1.200.1 | 10 | B.1.1.200.1, AN.1 | 33 |
| B.1.147 | 10 | B.1.147 | 0.84 |
| B.1.177.81 | 10 | B.1.177.81 | 1.8 |
| B.1.1.37 | 9 | B.1.1.37 | 0.42 |
| B.1.177.33 | 8 | B.1.177.33 | 4.5 |
| B.1.36 | 8 | B.1.36 | 0.8 |
| B.1.509 | 8 | B.1.509 | 2.3 |
| B.1.1.433 | 7 | B.1.1.433 | 7.8 |
| B.1.1.521 | 7 | B.1.1.521 | 19 |
| B.1.36.17 | 7 | B.1.36.17 | 1.2 |
| B.1.8 | 7 | B.1.8 | 1.8 |
| B.1.91 | 7 | B.1.91 | 1.6 |
| B.1.177.51 | 6 | B.1.177.51 | 20 |
| B.1.258.17 | 6 | B.1.258.17 | 1.4 |
| B.1.467 | 6 | B.1.467 | 33 |
| B.1.1.242 | 5 | B.1.1.242 | 35 |
| B.1.1.58 | 5 | B.1.1.58 | 14 |
| B.1.177.83 | 5 | B.1.177.83 | 7.8 |
| B.1.177.85 | 5 | B.1.177.85 | 11 |
| B.1.535 | 5 | B.1.535 | 0.59 |
| B.40 | 5 | B.40 | 0.23 |
| B.1.1.218 | 4 | B.1.1.218 | 5.2 |
| B.1.1.241 | 4 | B.1.1.241, AH | 4.2 |
| B.1.1.428 | 4 | B.1.1.428 | 50 |
| B.1.1.464 | 4 | B.1.1.464, AW | 1.2 |
| B.1.258.14 | 4 | B.1.258.14 | 11 |
| B.1.356 | 4 | B.1.356 | 0.82 |
| A | 3 | A | 0.15 |
| B.1.1.170 | 3 | B.1.1.170 | 2.6 |
| B.1.1.231.1 | 3 | B.1.1.231.1, AL.1 | 0.34 |
| B.1.1.297 | 3 | B.1.1.297, AG | 1.9 |
| B.1.1.317 | 3 | B.1.1.317, AS | 2.1 |
| B.1.1.371 | 3 | B.1.1.371 | 6.2 |
| B.1.177.52 | 3 | B.1.177.52, Y | 2.8 |
| B.1.177.53 | 3 | B.1.177.53, W | 3.6 |
| B.1.389 | 3 | B.1.389 | 1.5 |
| B.1.474 | 3 | B.1.474 | 14 |
| B.1.480 | 3 | B.1.480 | 4.3 |
| B.1.9.5 | 3 | B.1.9.5 | 11 |
| B.11 | 3 | B.11 | 1.8 |
| B.3 | 3 | B.3 | 0.37 |

| | | | |
|---|---|---|---|
| A.2 | 2 | A.2 | 0.22 |
| B.1.1.219 | 2 | B.1.1.219 | 1.6 |
| B.1.1.243 | 2 | B.1.1.243 | 4.2 |
| B.1.1.33 | 2 | B.1.1.33, N | 0.11 |
| B.1.1.44 | 2 | B.1.1.44 | 0.58 |
| B.1.1.50 | 2 | B.1.1.50 | 1.2 |
| B.1.160.28 | 2 | B.1.160.28 | 1.4 |
| B.1.177.15 | 2 | B.1.177.15, AA | 0.21 |
| B.1.177.32 | 2 | B.1.177.32 | 1.1 |
| B.1.177.53.1 | 2 | B.1.177.53.1, W.1 | 7.7 |
| B.1.177.55 | 2 | B.1.177.55 | 0.87 |
| B.1.177.60 | 2 | B.1.177.60, U | 2.5 |
| B.1.177.62 | 2 | B.1.177.62 | 6.2 |
| B.1.177.80 | 2 | B.1.177.80 | 17 |
| B.1.177.82 | 2 | B.1.177.82 | 0.63 |
| B.1.177.86 | 2 | B.1.177.86 | 2.1 |
| B.1.218 | 2 | B.1.218 | 6.5 |
| B.1.408 | 2 | B.1.408 | 3.5 |
| B.1.416 | 2 | B.1.416 | 0.94 |
| B.1.523 | 2 | B.1.523 | 0.88 |
| B.1.9.4 | 2 | B.1.9.4 | 12 |
| B.28 | 2 | B.28 | 0.57 |
| B.4 | 2 | B.4 | 0.54 |
| B.58 | 2 | B.58 | 2.2 |
| B.59 | 2 | B.59 | 1.3 |
| A.5 | 1 | A.5 | 0.21 |
| B.1.1.1.30 | 1 | B.1.1.1.30, C.30 | 0.19 |
| B.1.1.142 | 1 | B.1.1.142 | 6 |
| B.1.1.145 | 1 | B.1.1.145 | 4.5 |
| B.1.1.198 | 1 | B.1.1.198 | 0.18 |
| B.1.1.221 | 1 | B.1.1.221 | 1.2 |
| B.1.1.266 | 1 | B.1.1.266 | 4.9 |
| B.1.1.28 | 1 | B.1.1.28, P | 0.066 |
| B.1.1.294 | 1 | B.1.1.294, M | 0.28 |
| B.1.1.294.2 | 1 | B.1.1.294.2, M.2 | 50 |
| B.1.1.315 | 1 | B.1.1.315, AD | 1.4 |
| B.1.1.331 | 1 | B.1.1.331 | 2.4 |
| B.1.1.336 | 1 | B.1.1.336 | 7.1 |
| B.1.1.355 | 1 | B.1.1.355 | 2.5 |
| B.1.1.369 | 1 | B.1.1.369 | 0.046 |
| B.1.1.406 | 1 | B.1.1.406 | 3.1 |
| B.1.1.409 | 1 | B.1.1.409 | 0.83 |

| | | | |
|---|---|---|---|
| B.1.1.519 | 1 | B.1.1.519 | 2.6 |
| B.1.1.71 | 1 | B.1.1.71 | 1.3 |
| B.1.12 | 1 | B.1.12 | 0.89 |
| B.1.127 | 1 | B.1.127 | 0.53 |
| B.1.149 | 1 | B.1.149 | 2.7 |
| B.1.177.31 | 1 | B.1.177.31 | 50 |
| B.1.177.50.1 | 1 | B.1.177.50.1, Z.1 | 0.5 |
| B.1.177.53.3 | 1 | B.1.177.53.3, W.3 | 0.65 |
| B.1.177.6 | 1 | B.1.177.6 | 0.19 |
| B.1.177.7 | 1 | B.1.177.7 | 0.03 |
| B.1.177.72 | 1 | B.1.177.72 | 1.8 |
| B.1.2 | 1 | B.1.2 | 0.0053 |
| B.1.213 | 1 | B.1.213 | 4.2 |
| B.1.220 | 1 | B.1.220 | 1.2 |
| B.1.221.1 | 1 | B.1.221.1 | 0.34 |
| B.1.229 | 1 | B.1.229 | 1.1 |
| B.1.258.4 | 1 | B.1.258.4 | 0.46 |
| B.1.258.7 | 1 | B.1.258.7 | 0.27 |
| B.1.258.9 | 1 | B.1.258.9 | 0.65 |
| B.1.36.22 | 1 | B.1.36.22 | 0.24 |
| B.1.36.24 | 1 | B.1.36.24 | 4.5 |
| B.1.36.35 | 1 | B.1.36.35 | 2.1 |
| B.1.397 | 1 | B.1.397 | 0.86 |
| B.1.398 | 1 | B.1.398 | 1.4 |
| B.1.400 | 1 | B.1.400 | 0.099 |
| B.1.406 | 1 | B.1.406 | 1.9 |
| B.1.415 | 1 | B.1.415 | 1.4 |
| B.1.513 | 1 | B.1.513 | 1.4 |
| B.1.520 | 1 | B.1.520 | 0.1 |
| B.1.540 | 1 | B.1.540 | 1.6 |
| B.1.88.1 | 1 | B.1.88.1 | 0.6 |
| B.39 | 1 | B.39 | 0.26 |
| B.55 | 1 | B.55 | 0.55 |
| B.6 | 1 | B.6 | 0.14 |
| None | 1 | None | 1 |

**Table S2.** Top 20 largest SARS-CoV-2 sequencing data contributors to GISAID in 2020 by submitting lab.

| Submitting lab | Countries represented (ISO codes) | Number of sequences |
|---|---|---|
| Wellcome Sanger Institute for the COVID-19 Genomics UK (COG-UK) Consortium | GBR | 96441 |
| COVID-19 Genomics UK (COG-UK) Consortium | GBR | 71371 |
| Albertsen Lab, Department of Chemistry and Bioscience, Aalborg University, Denmark | DNK | 27936 |
| Houston Methodist Hospital | USA | 27409 |
| Pathogen Genomics Center, National Institute of Infectious Diseases | JPN; MMR | 19708 |
| Department of Biosystems Science and Engineering, ETH Zürich | CHE | 11357 |
| MDU-PHL | AUS; TLS | 10459 |
| TGen North | USA | 9491 |
| Wyoming Public Health Laboratory | USA | 9172 |
| Aalborg University | DNK | 8439 |
| SeqCOVID-SPAIN consortium/IBV(CSIC) | ESP | 8279 |
| Chan-Zuckerberg Biohub | USA | 7803 |
| BCCDC Public Health Laboratory | CAN | 7646 |
| Laboratoire de santé publique du Québec | CAN | 6914 |
| Andersen lab at Scripps Research | JOR; MEX; USA | 6258 |
| Utah Public Health Laboratory | USA | 5925 |
| MEPHI, Aix Marseille University | FRA | 5617 |
| Respiratory Virus Unit, Microbiology Services Colindale, Public Health England | GBR; UKR | 5142 |
| deCODE genetics | ISL | 5005 |
| Erasmus Medical Center | BEL; BHR; LUX; NLD; SUR | 4594 |

**Table S3.** Sampling proportion change-points for the phylodynamic analysis on Swiss data. The sampling proportion was modeled as a piecewise-constant function in time, with the following change-points motivated by major shifts in the testing regime or genome sequencing intensity in Switzerland.

| Start date | Description |
|---|---|
| 23 April 2020 | All symptomatic individuals can get tested |
| 25 June 2020 | Government pays for tests for symptomatic individuals |
| 14 September 2020 | Genome sequencing << 5% of confirmed cases |
| 28 September 2020 | Number of tests conducted and % positivity dramatically increase, genome sequencing also increases |
| 19 October 2020 | Genome sequencing << 5% of confirmed cases again |
| 11 November 2020 | Genome sequencing increases again |
